# Clinical outcomes and risk factors for COVID-19 among migrant populations in high-income countries: a systematic review

**DOI:** 10.1101/2020.12.21.20248475

**Authors:** Sally E Hayward, Anna Deal, Cherie Cheng, Alison F Crawshaw, Miriam Orcutt, Tushna F Vandrevala, Marie Norredam, Manuel Carballo, Yusuf Ciftci, Ana Requena-Mendez, Chris Greenaway, Jessica Carter, Felicity Knights, Anushka Mehrotra, Farah Seedat, Kayvan Bozorgmehr, Apostolos Veizis, Ines Campos-Matos, Fatima Wurie, Teymur Noori, Martin McKee, Bernadette N Kumar, Sally Hargreaves, the ESCMID Study Group for Infections in Travellers and Migrants (ESGITM)

**Affiliations:** Institute for Infection and Immunity, St George’s University of London; Institute for Global Health, University College London; Faculty of Business and Social Sciences, Kingston University; Danish Research Centre for Migration, Ethnicity and Health, University of Copenhagen; International Centre for Migration, Health, and Development, Geneva, Switzerland; Doctors of the World UK, London, UK; Department of Medicine-Solna, Karolinska Institutet, Sweden; Barcelona Institute for Global Health (ISGlobal-University of Barcelona), Spain; Department of Medicine, McGill University; Department of Population Medicine and Health and Health Services Research, School of Public Health, Bielefeld University; Section for Health Equity Studies & Migration, Heidelberg University Hospital; Medecins Sans Frontieres Greece, Athens, Greece; Public Health England; Public Health England; and UCL Collaborative Centre for Inclusion Health, London, UK; Public Health England; and UCL Research Department of Epidemiology and Public Health; European Centre for Disease Prevention and Control, Stockholm, Sweden; Norwegian Institute of Public Health; Faculty of Public Health and Policy, London School of Hygiene and Tropical Medicine

## Abstract

**Background:** Migrants, including refugees, asylum seekers, labour migrants, and undocumented migrants, now constitute a considerable proportion of most high-income countries’ populations, including their skilled and unskilled workforces. Migrants may be at increased risk of COVID-19 due to their health and social circumstances, yet the extent to which they are being affected and their predisposing risk factors are not clearly understood. We did a systematic review to assess clinical outcomes of COVID-19 in migrant populations (cases, hospitalisations, deaths), indirect health and social impacts, and to determine key risk factors.

**Methods:** We did a systematic review following PRISMA guidelines, registered with PROSPERO (CRD42020222135). We searched databases including PubMed, Global Health, Scopus, CINAHL, and pre-print databases (medRxiv) via the WHO Global Research on COVID-19 database to Nov 18, 2020 for peer-reviewed and grey literature pertaining to migrants (defined as foreign born) and COVID-19 in 82 high-income countries. We used our international networks to source national datasets and grey literature. Data were extracted on our primary outcomes (cases, hospitalisations, deaths) and we evaluated secondary outcomes on indirect health and social impacts, and risk factors, using narrative synthesis.

**Results:** 3016 data sources were screened with 158 from 15 countries included in the analysis (35 data sources for primary outcomes: cases [21], hospitalisations [4]; deaths [15]; 123 for secondary outcomes). We found that migrants are at increased risk of infection and are disproportionately represented among COVID-19 cases. Available datasets suggest a similarly disproportionate representation of migrants in reported COVID-19 deaths, as well as increased all-cause mortality in migrants in some countries in 2020. Undocumented migrants, migrant health and care workers, and migrants housed in camps and labour compounds may have been especially affected. In general, migrants have higher levels of many risk factors and vulnerabilities relevant to COVID-19, including increased exposure to SARS-CoV-2 due to high-risk occupations and overcrowded accommodation, and barriers to health care including inadequate information, language barriers, and reduced entitlement to healthcare coverage related to their immigration status.

**Conclusions:** Migrants in high-income countries are at high risk of exposure to, and infection with, COVID-19. These data are of immediate relevance to national public health responses to the pandemic and should inform policymaking on strategies for reducing transmission of COVID-19 in this population. Robust data on testing uptake and clinical outcomes in migrants, and barriers and facilitators to COVID-19 vaccination, are urgently needed, alongside strengthening engagement with diverse migrant groups.

## Introduction

The COVID-19 pandemic has highlighted the vast ethnic, social, economic and cultural diversity that has come to characterise contemporary high-income countries (HICs), and has served as a reminder of the growing rate of population movement between, as well as within, countries and the new public health opportunities and challenges this is presenting. One of these challenges is the scale of health and social disparities associated with this diversity, with profound consequences for some ethnic minority groups (1). Data from several countries have revealed a much greater risk of infection and adverse outcomes from COVID-19 among Black, Asian, and Minority Ethnic [BAME] groups, South/East Asian, Black Americans, Hispanics, Latinos, racialised groups, people of colour, and indigenous groups compared to the native white population in the same countries (2). These adverse outcomes are likely the result of a complex interaction of socioeconomic disadvantage influencing exposure to SARS-CoV-2 and underlying health status, that predisposes to severe illness (3, 4), leading to calls to address the root causes of these inequalities now and in the future.

Although a picture is emerging, there is not yet a comprehensive overview of the extent to which migrants (defined as foreign-born) – including refugees, asylum seekers, labour migrants, and undocumented migrants living temporarily or permanently in different HICs – have been impacted by COVID-19, and their specific risk factors. Prior to the COVID-19 pandemic, global migration was at its highest level on record, with 1 billion people on the move around the world, and with HICs receiving unprecedented numbers of people seeking human security either through political asylum and/or work opportunities (5). Most of the relatively few health datasets with information on ethnicity currently used to monitor COVID-19 reflect what information is already recorded by healthcare systems (which is highly variable across countries and regions). For the most part these fail to capture migration status, combining those born in the host countries to families that may have been in the country for several generations with more recent migrants, thus failing to reflect the health dynamics of contemporary migration. Although more recently arrived migrants predominantly from low- and middle-income countries, are typically considered to be young and healthy on arrival (6), and may share many of the characteristics of “older” generation ethnic minorities and their offspring, they may also present a unique spectrum of health and social risk factors for COVID-19 exposure and infection that to date has been poorly defined.

In many countries, migrants make up a significant proportion of front-line workers who may have a greater exposure to COVID-19, in sectors witnessing a disproportionate impact of COVID-19 infections (7). There are, in addition, tens of thousands of migrants in HICs who are being housed in camps, detention centres, and labour dormitories or compounds, all of which are considered high-risk environments for COVID-19. Recent analyses suggest that countries and regions with large migrant populations (including US, Italy, Spain, France, and the UK) should ensure they are better considered in public health responses (8, 9).

In order to develop a more targeted and inclusive public health response a better understanding of the impact that COVID-19 is having specifically on migrant populations is critically needed. We therefore did a systematic review to explore and assess what is currently known about clinical outcomes of COVID-19 (cases, hospitalisations, deaths), indirect health and social impacts, and to identify key risk factors and vulnerabilities in migrant populations.

## Methods

### Search strategy

We undertook a systematic review in line with the Preferred Reporting Items for Systematic Reviews and Meta-Analyses (PRISMA) guidelines (10), and registered with PROSPERO (CRD42020222135). We searched the following databases: Embase, Web of Science, Oxford Academic Journals, PubMed NIH, Clinical Trials, China CDC MMWR, CDC reports, ProQuest Central (Proquest), CINAHL, Africa Wide Information (Ebsco), Scopus, PsycInfo, CAB Abstracts, Global Health, J Stage, Science Direct, Wiley Online Journals, JAMA Network, British Medical Journal, Mary Ann Liebert, New England Journal of Medicine, Sage Publications, Taylor and Francis Online, Springer Link, Biomed Central, MDPI, ASM, PLOS, The Lancet, Cell Press, and pre-print sites chemRxiv, SSRNbioRxiv, and medRxiv facilitated through the WHO Global Research on COVID-19 database from inception to 18/11/2020 (https://search.bvsalud.org/global-literature-on-novel-coronavirus-2019-ncov/). The latter is a daily-updated, multilingual resource of all the global literature (peer-reviewed literature, pre-prints and grey literature) pertaining to COVID-19. We used a broad search strategy encompassing terms related to ethnicity and migrants, to source specific information pertaining to migrants (Appendix 1).

Records were imported into EndNote, and duplicates deleted. Title/abstract and full-text screening were carried out by two reviewers using Rayyan QCRI (11). A snowballing method was used to follow up potentially relevant articles cited in included papers. Grey literature sources were also hand-searched. Our international networks were used to directly engage migrant health experts in key countries, who were specifically approached to source country-level public health data (via the Ministry of Health and public health statistics) and other grey literature.

### Selection criteria and primary/secondary outcomes

We included any data pertaining to our selected primary and secondary outcomes on migrant populations from 82 World Bank HICs (countries listed in Appendix 2). Migrants were defined as foreign-born individuals, born outside of the country in which they are resident. Primary outcomes were clinical outcomes of COVID-19 in migrant populations (cases, hospitalisation, deaths). Secondary outcomes included indirect health and social impacts, and risk factors and vulnerabilities (co-morbidities, health behaviours and systemic factors, social and cultural factors, and occupation).

No restrictions were imposed on study design because our preliminary scoping review revealed that in this rapidly evolving field important data were often embedded into letters, editorials, and grey literature as well as primary research studies and national statistics. We imposed no language restrictions and information was translated where required. Studies were excluded if it was not possible to determine whether individual(s) in the population studied were migrants, based on the stated criteria, and where data were collected outside of the countries listed or did not directly relate to COVID-19 outcomes, impacts and risk factors. We excluded all mass media reports.

### Data extraction, critical appraisal and synthesis

Abstracts were screened and data were extracted in duplicate at each stage, involving three researchers (CC, SEH, SH). Records and data were managed through EndNote and Excel databases prepared by the principal reviewers. The quality of studies was assessed by two reviewers (AD, CC), using Joanna Briggs Institute critical appraisal tools (checklists for cohort studies, qualitative research, prevalence studies, cross-sectional studies, case series or text and opinion checklists, as appropriate for the individual study design) (12). Quality scores were calculated as a total out of the maximum number of applicable questions and converted into percentages. Studies with a score of 80-100% were considered high quality, 60-79% medium quality and 0-59% low quality. Data sources were not excluded based on study quality, but information on quality contributed to the meta-synthesis and discussion. Only original research was appraised for both primary and secondary outcomes, as the appraisal tools are specific for study designs and thus are not applicable to sources such as commentaries. Critical appraisals were only performed for literature in English, French or Spanish, due to the language restrictions of the critical appraisal team.

For the primary outcomes we included only primary data sources; the heterogeneity of study designs and populations precluded meta-analysis. For the secondary outcomes we included primary data and data from other sources, which was collated and assessed using narrative synthesis.

## Results

Initial searches of databases and for grey literature identified 3016 records to screen; 158 of which were included in the final analysis (35 for primary outcomes, 123 for secondary outcomes) (Figure 1). Supplementary Table 1 details the characteristics of all included data sources.

**Figure 1.**
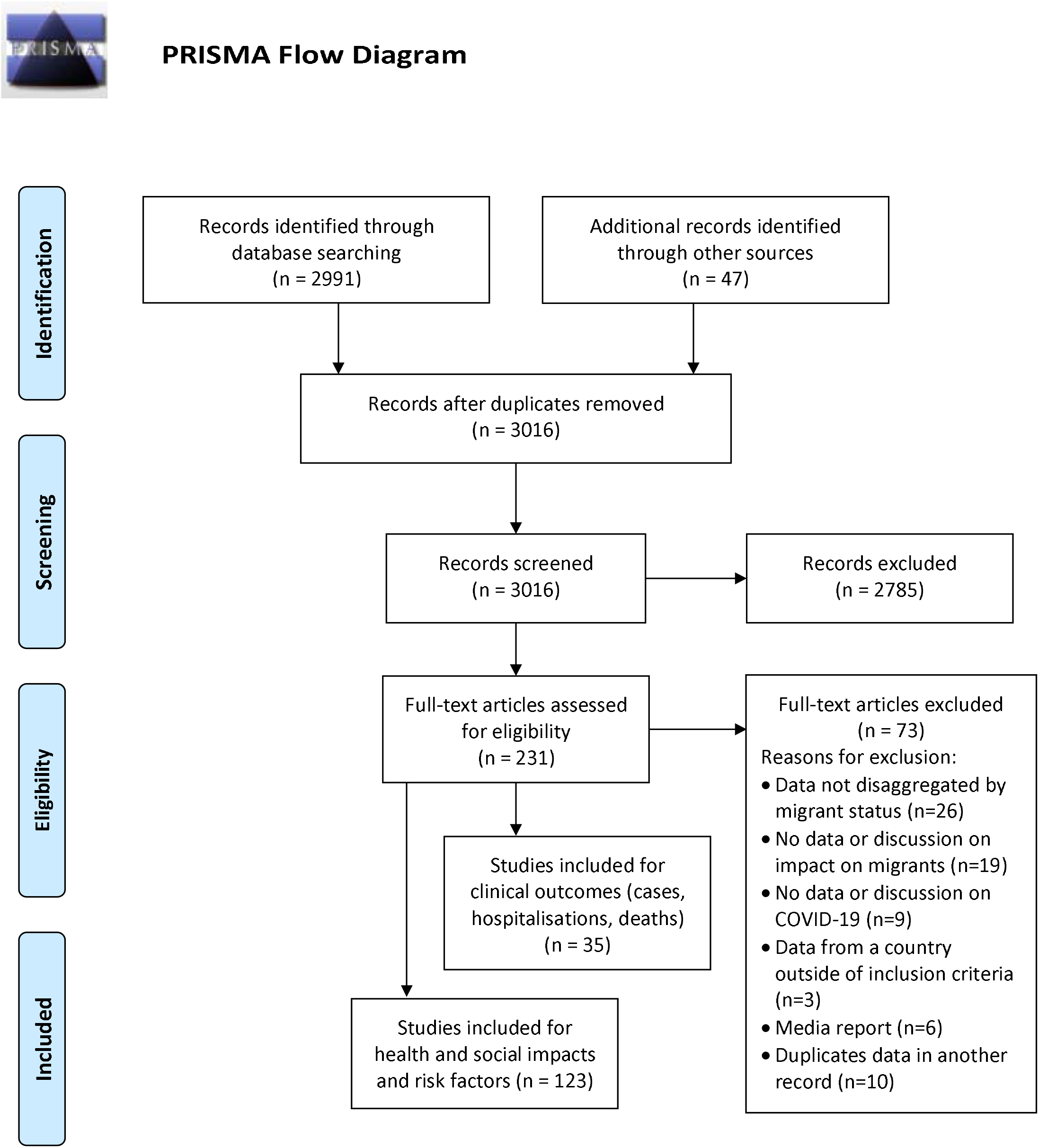
PRISMA flow diagram of included data sources.

We found 35 data sources reporting on our primary clinical outcomes in migrants, including 21 on cases (13–32), 4 on hospitalisations (20, 33–35), and 15 on mortality (3, 20, 33–45). This includes data from Sweden (6 records), Italy (4), the United States (3), Canada (2), Denmark (2), Spain (2), the UK (2), France (2), Kuwait (2), Singapore (2), Norway (2), Germany (2), the Netherlands (1), Greece (1), Saudi Arabia (1) and across the EU/EEA/UK (1). Sources include peer-reviewed journal articles (13 records), pre-prints (3), national statistics (10), and other grey literature (9). A total of 59 studies were subjected to critical appraisal, including 22 primary outcomes and 37 secondary outcomes. Literature ranged in quality, with 28 studies fitting the criteria for high quality studies (80-100%), 19 for medium quality (60-79%) and 12 for low quality (0-59%). The average quality appraisal score was 73.6%, with reports included in the primary outcomes having a slightly higher quality score on average than those included in the secondary outcomes (74.9% and 72.9%, respectively). An additional 123 studies reported on indirect impacts of the pandemic on migrants and/or on risk factors for COVID-19 in migrants.

### Clinical outcomes

Table 1 summarises included studies on the primary outcome (cases, hospitalisation, deaths).

**Table 1.** Data sources included in primary outcome data, clinical outcomes (cases, hospitalisations, deaths)

| Authors* | Location | Population | Study design | Publication type | Study period | Sample size | Methods | Key results | Quality appraisal score (%) |
| --- | --- | --- | --- | --- | --- | --- | --- | --- | --- |
| *COVID-19 cases* |  |  |  |  |  |  |  |  |  |
| Guttman (13) | Ontario, Canada | Migrants and refugees | Population-based case/testing data | Grey | To 13 June | 624,386 tested | Rates of COVID-19 testing and percentage positivity in migrants, and relationship with socioeconomic factors | Migrants accounted for 43.5% of all COVID-19 cases but make up just over 25% of the population; migrants had lower rates of testing but a higher percentage positivity in those tested (refugees 10.4% positive, other migrants 7.6%, and Canadian-born 2.6%) | 6/10 (60) |
| Sundaram (14) | Ontario, Canada | Migrants | Spatial comparison | Pre-print | 1 March to 20 June | 25,050 diagnoses | Association between COVID-19 diagnosis and percentage of migrants by area | Living in an area with a greater percentage of immigrants was positively associated with rate of COVID-19 diagnosis | 8/8 (100) |
| Kim (15) | Washington DC, | Non-English | Case/testing data | Peer-reviewed | 29 February to | 562,242 tested | Rates of COVID-19 | Non-English speakers were tested less | 6/8 (75) |
|  | USA | speakers |  | publication | 31 May |  | testing and percentage positivity in non-English speakers versus English speakers | frequently for COVID-19 (4.7% vs 5.6%) but were more likely to receive a positive result (18.6% vs 4.0%) |  |
| Norwegian Institute of Public Health (16) | Norway | Migrants | Population-based case data | National statistics | To 4 May | 7,847 cases | Percentage of reported cases that are among the foreign-born | Migrants made up 19% of reported cases in week 12 and 42% in week 18 | N/A |
| Norwegian Institute of Public Health (17) | Norway | Migrants | Population-based case data | National statistics | To 5 Nov | - | Incidence rate among migrants by country of origin versus those born in Norway | Until 1 July, incidence among the Somali-born was very high, but in autumn the risk has increased for migrants from Pakistan, Iraq, Afghanistan, Serbia and Turkey | N/A |
| Swedish Public Health Agency (18) | Sweden | Migrants | Population-based case data | National statistics | 13 March to 7 May | - | Incidence of COVID-19 by country of birth | 32% of cases were immigrants, despite making up 19% of the population; highest incidence among those born in Turkey, Ethiopia, and Somalia | N/A |
| Statens Serum Institut (19) | Denmark | Migrants and their children | Population-based case data | National statistics | 29 April to 6 May | - | Case, testing and incidence data, comparing migrants and their descendants with non-migrants | Non-Western migrants and their native-born children accounted for 18% of cases, despite making up 9% of the population | N/A |
| Statens Serum Institut (20) | Denmark | Migrants and their children | Population-based data on cases, hospitalisation and mortality | National statistics | 29 April to 7 September | - | Case, testing incidence, hospitalisation and mortality data, comparing migrants and their descendants with non-migrants | Non-Western migrants and their native-born children accounted for 26% of cases, and 15% of COVID-19 hospital admissions, despite making up 9% of the population | N/A |
| Guijarro (21) | Alcorcón, Spain | Migrants | Population-based cohort study | Pre-print | 1 February to 25 April | 152,018 residents | Incidence of COVID-19 in migrants versus Spaniards and relative risk by region of origin | Crude incidence of COVID-19 among migrants was higher than among Spaniards, at 8.71 and 6.51 per 1000 inhabitants respectively (p<0.001) | 8/8 (100) |
| Grilli (22) | Reggio Emilia, Italy | Migrants | Population-based case data | Peer-reviewed publication | 6 March to 26 March | 2635 tested | Odds of COVID-19 infection and being tested in migrants versus Italians | Immigrants and Italians had a similar prevalence of infection (OR 0.99, 95% CI 0.82-1.20) and similar probability of being tested (OR 0.93, 95% CI 0.81-1.10) | N/A |
| Strully (23) | USA | Migrants | Spatial comparison | Peer-reviewed publication | To 28 May | - | Association of proportion of migrants living in a region with COVID-19 case rates | Percentage of foreign-born residents was positively associated with COVID-19 case rate (fully adjusted IRR = 1.106, 95% CI 1.074-1.139, p<0.01) at county level | 8/8 (100) |
| Jaqueti Aroca (24) | Madrid, Spain | Migrants | Case/testing data | Peer-reviewed publication | To the second week of April | 1,781 patients | Percentage positivity among foreign-born versus | No significant difference in percentage positivity between migrants and Spaniards (OR 1.08, 95% CI 0.95-1.24), but those | 8/10 (80) |
|  |  |  |  |  |  |  | Spaniards in hospitals | from Latin America are at higher risk; only 12.5% of positive migrants were >65 years versus 56.9% Spaniards |  |
| Chew (25) | Singapore | Migrant workers | Case/testing data and clinical evaluation | Peer-reviewed publication | 11 to 19 April | 5,977 migrant workers | Review of data from an outbreak investigation among migrant workers in a dormitory, including test positivity and clinical parameters | 1832 of 5977 migrant workers were symptomatic, of which 1264 (69%) were positive for COVID-19, corresponding to 21% of the cohort | 6/10 (60) |
| Alkhamis (26) | Kuwait | Migrant workers | Population-based case data | Peer-reviewed journal | 23 February to 7 May | 5988 cases | Modelling pandemic progression (spatiotemporal cluster analysis) in Kuwait citizens/ | 78.8% of COVID-19 cases were in migrant workers, 40.1% of which were of Indian nationality; significant spreading events among migrant workers | 6/10 (60) |
|  |  |  |  |  |  |  | residents and migrant workers |  |  |
| Openshaw (27) | USA | Migrants in detention centres | Viewpoint (in | Peer-reviewed publication | To May | - | Reports COVID-19 cases at ICE detention centres | Over 1200 confirmed COVID-19 cases across 52 facilities run by ICE | 6/6 (100) |
| ECDC (28) | EU/EEA and UK | Migrants in detention centres | Report reporting cases | Grey | To June | - | Reports COVID-19 cases at detention centres in Europe | Reported outbreaks in detention centres in Germany and Portugal | 6/6 (100) |
| Ministry of Health (29) | Singapore | Migrant workers | Population-based case data | National statistics | 18 Nov | 5,704,000 (population ) | Surveillance of new confirmed cases in the past 14 days, including proportion in dorm residents | 54,502 (95.7%) of 58,135 all in-country cases of COVID-19 were in migrants residing in dormitories | 2/10 (20) |
| Ministry of | Saudi | Migrants | Population- | National | To 7 May | - | Routine | 75% of all people in- | N/A |
| Health (30) | Arabia |  | based case data | statistics |  |  | surveillance | country who have tested positive for COVID-19 were migrants |  |
| Greek Ministry of Health (47) | Greece | Migrants and refugees | Hospital-based case data | National statistics | To 16 September | - | Routine surveillance | Almost half of COVID-19 patients hospitalised in Attica are refugees from camps/hosting sites or destitute migrants | N/A |
| Buda (31) | Germany | Refugees | Outbreaks case data | National statistics | To 11 August | - | Collation of outbreak reports, assessing the proportion in refugee centres (vs other settings such as nursing homes) | 2.5% of notified outbreaks (199 of 7864) were reported in refugee centres, comprising 7.5% (n=4,146) of all notified cases during outbreaks (n=55,141) | N/A |
| Bozorgmehr (32) | Germany | Refugees in reception/accommodation centres | Outbreaks case data | Grey | To 22 May | 9785 refugees | Meta-analysis of media reports in Germany to identify pooled cumulative incidence rate in refugee reception/ accommodation centres | Identified 42 outbreaks in 11 federal states, with 1769 confirmed cases; IR of 17.0% (95% CI 12.0 to 23.0, $I^2 = 98.3\%$) | N/A |
| *COVID-19 hospitalisation* |  |  |  |  |  |  |  |  |  |
| Giorgi Rossi (33) | Reggio Emilia, Italy | Migrants | Population-based cohort study | Peer-reviewed publication | 27 February to 2 April | 2,653 tested | COVID-19 incidence, hospitalisation and death in migrants versus Italians | Immigrants had a higher risk of hospitalisation (HR 1.3, 95% CI 0.99-1.81) than Italians | 7/10 (70) |
| Hamadah (34) | Kuwait | Migrants | Hospital-based cohort study | Peer-reviewed publication | 24 February to 20 April | 1,123 patients | Comparison of ICU admission, ARDS, pneumonia and mortality in migrants and non-migrants | Migrants had increased odds of death or ICU admission (OR 2.14, 95% CI 1.12-4.32), ARDS (OR 2.44, 95%CI 1.23-5.09) and pneumonia (OR 2.24, 95% CI 1.27-4.12) | 8/8 (100) |
| Fabiani (35) | Italy | Migrants | Population-based clinical and | Pre-print | 20 Feb to 19 July | 213,180 cases | Comparison of case fatality rate and rate | Non-Italian cases were diagnosed at a later date than Italian cases and | 8/8 (100) |
|  |  |  | mortality data |  |  |  | of admission to hospital and ICU between migrants versus Italians | were more likely to be hospitalised (ARR=1.39, 95% CI 1.33- 1.44) and admitted to an ICU (ARR=1.19, 95% CI 1.07- 1.32) |  |
| *COVID-19 mortality* |  |  |  |  |  |  |  |  |  |
| Canevelli (36) | Italy | Migrants | Temporal comparison | Peer-reviewed publication | 21 February to 29 April | 2,687 deceased cases | Comparison of proportion of migrants in COVID-19 deaths versus all-cause mortality in 2018 | The proportion of migrants and non-migrants among COVID-related deaths (2.5% and 97.5% respectively) was similar to the estimated 2018 all-cause mortality rates (2.6% and 97.4%); but migrants were younger at the time of death versus non-migrants (71.1, SD 13.1 vs 78.3, SD 10.8, p<0.001) | 9/10 (90) |
| Public Health England (3) | England | Migrants | Temporal comparison | Grey | 21 March to 8 May | - | Comparison of all-cause mortality in 2020 versus | Deaths in 2020 were over 3 times higher than 2014-2018 for those from Central and | 8/8 (100) |
|  |  |  |  |  |  |  | 2014-2018 in migrants and UK-born | Western Africa, the Caribbean, South East Asia, Middle East, and South and Eastern Africa, versus 1.7 times higher overall in England |  |
| Papon (37) | France | Migrants | Temporal comparison | Grey | March to April | - | Comparison of proportion of migrants in registered deaths in 2020 versus 2014-2019 | The foreign-born represented 15% of registered deaths in March and April 2020 versus 13% in March and April 2019 | 6/10 (60) |
| Observatoire Regional de Sante Ile de France (38) | Paris, France | Migrants | Spatial comparison | Grey | March 2020 | Not stated | Mortality (daily deaths) by Parisian departments (areas) compared with sociodemographic characteristics of the department. | Eg. Seine-Saint-Denis, a district in the north of Paris where 30% of the population is an immigrant, had a 188% mortality increase compared with 2019 versus a 96% increase in Paris as a whole | 1/10 (10) |
| Kunst (39) | Netherlands | Migrants and their children | Temporal comparison | National statistics | March to April | - | Comparison of mortality in March-April versus in the preceding weeks, adjusted for seasonal factors, in migrants versus Dutch | Mortality was 47% higher than expected for immigrants from non-Western countries and their children, 49% higher for immigrants from Western countries and their children, and 38% higher for the native-born with Dutch parents | N/A |
| Hansson (40) | Sweden | Migrants | Temporal comparison | Peer-reviewed publication | February to May | - | Comparison of all-cause mortality in 2020 versus 2016-2019 by region of birth | Among middle-aged (40-64 years) and older (>65 years) people born in Syria, Iraq and Somalia excess mortality was ~220%; among those born in Sweden, the EU, the Nordic countries or North America, excess mortality among those >65 was 19% and among the middle aged was 1% | N/A |
| Calderón-Larrañaga | Stockholm, | Migrants | Spatial/temporal | Peer-reviewed | 6-12 April | 2,379,792 residents | Comparison of excess | Areas with the lowest tercile share of Swedish- | 6/10 (60) |
| (41) | Sweden |  | comparison | publication |  |  | mortality compared with previous 5 years in areas according to share of migrants | born had 178% excess mortality compared with the previous 5 years |  |
| Drefahl (42) | Sweden | Migrants | Individual-level survival analysis | Peer-reviewed publication | To 7 May | 1,189,484 py (17,181 deaths) | Risk of death from COVID-19 in individual-level data according to migrant status and region of origin | Immigrants from LMICs from the Middle East and North Africa showed increased mortality among men (HR 3.13, 95% CI 2.51-3.90) and women (HR 2.09, 95% CI 1.52-2.89) as compared to the Swedish-born | 8/10 (80) |
| Rostila (43) | Stockholm, Sweden | Migrants | Population-based cohort study | Grey | 31 Jan to 4 May | 1,778,670 individuals | Risk of death from COVID-19 in individual-level data according to migrant status | Migrants from Middle Eastern countries (RR 3.2, 95% CI 2.6-3.8), Africa (RR 3.0, 95% CI 2.2-4.3) and the Nordic countries (RR 1.5, 95% CI 1.2-1.8) had higher | 9/11 (82) |
|  |  |  |  |  |  |  | and region of origin | COVID-19 mortality versus the Swedish-born |  |
| Centrum for epidemiologi och samhällsmedicin (44) | Stockholm, Sweden | Migrants | Individual-level survival analysis | Grey | To 30 June | - | Risk of death from COVID-19 in individual-level data according to country of birth, among those aged 25 years and older | Migrants from Somalia (HR 12.39, 95% CI 7.93-19.36), Lebanon (HR 6.19, 95% CI 3.41-11.24), and Syria (HR 6.14, 95% CI 4.28-8.80) show increased risk of death compared with Swedish-born, adjusted for age and sex | N/A |
| Cook (45) | UK | Migrant healthcare workers (HCWs) | Characterisation of reported HCW deaths | Grey | To 22 April | 106 HCWs | Proportion of UK healthcare workers who died who were born outside the UK | Of 106 UK healthcare workers who died up until 22 April 2020, at least 56 (53%) were born outside the UK | 4/10 (40) |

*\*Where papers report on multiple outcomes (cases, hospitalisations, deaths) papers are included under the first relevant sub-heading only*

### COVID-19 cases

Data that disaggregate COVID-19 incidence and testing uptake by migrant status indicate that migrants account for a disproportionate number of COVID-19 infections despite low rates of testing. In Ontario, Canada, immigrants make up just over 25% of the population, but accounted for 43.5% of all COVID-19 cases up until 13 June (13). Immigrants had lower rates of testing but there was a higher percentage of positive cases in those tested. Refugees had the highest percentage positivity, at 10.4%, compared with 7.6% among other immigrants, and 2.6% in the Canadian-born. Migrants and refugees from Central, Western and East Africa, South America, the Caribbean, Southeast Asia and South Asia showed the highest rates of positive cases for COVID-19 (13). Among all women who tested positive, 36% were employed as healthcare workers (immigrants and refugees made up 45% of these positive healthcare workers): 55% of positive cases were among female migrants in the economic caregiver categories, including 53% among those from the Philippines, 64% from Jamaica and 76% from Nigeria (13). In another study, living in an area of Ontario with a greater percentage of recently arrived migrants was significantly positively associated with an increased rate of COVID-19 diagnoses (14).

In the US, a study that reports using language as a surrogate for immigration status (in the absence of routine data collection on migrant status) found that non-English speakers were tested less frequently for COVID-19 (29 February to 31 May) (4.7% [95% CI 4.5%-4.9%] vs 5.6% [95% CI 5.6%-5.7%]), with variations across language groups, but were more likely to test positive (18.6% [95% CI 16.8%-20.4%] vs 4.0% [95% CI 3.8%-4.2%]) (15). Fewer years of formal education and a lack of English or French language ability at the time of immigration was associated with lower testing rates and higher percentage positivity among recent adult migrants in Ontario, Canada (13).

In Norway, migrants made up 19% of all reported cases in the week starting 16 March, rising to 42% in the week starting 27 April (16). While incidence among the Somali-born was very high until 1 July, in the autumn the risk increased for migrants from Pakistan, Iraq, Afghanistan, Serbia and Turkey (17). Similarly in Sweden, during the first peak of the pandemic (13 March to 7 May), 32% of positive cases were immigrants, despite making up only 19% of the population (18). The incidence of COVID-19 was highest among migrants from Turkey (753 per 100,000), followed by Ethiopia (742 per 100,000) and Somalia (660 per 100,000). This compares with an incidence of 189 per 100,000 for non-migrants who were born in Sweden for the same time-period (18).

In Denmark, non-Western migrants and their native-born children accounted for 18% of cases (29 April to 6 May), which was double their share in the Danish population (19). In an later update (7 September), this had risen substantially to migrants accounting for 26% of cases (20). Among non-Western migrants, the COVID-19 incidence rate was 315 per 100,000 compared with 240 per 100,000 for non-Western descendants and 128 per 100,000 among ethnic Danes (29 April to 6 May) (19). Particularly high incidence rates were seen among migrants from Morocco, Pakistan, Somalia and Turkey (19, 20).

In Alcorcón, Spain (to 25 April) the crude incidence rate of COVID-19 among migrants was higher than among the host Spanish populations, at 8.71 and 6.51 per 1000 inhabitants respectively (p<0.001) (21). The relative risk for COVID-19 was elevated in migrants from sub-Saharan Africa (RR 3.66, 95% CI 1.42-9.41; p=0.007), Caribbean (RR 6.35, 95% CI 3.83-10.55; p<0.001), and Latin America (RR 6.92, 95% CI 4.49-10.67; p<0.001) but not from other regions (21). Data from a hospital in Madrid up to the second week of April showed no significant differences between migrants and host population in terms of COVID-19 positivity among those tested (52.5% [136/259] vs 51.4% [782/1522]). There was also no difference in testing rate (odds ratio [OR] 1.08 95% CI 0.95-1.24) between migrants and the host population; only 12.5% of COVID-19 positive migrants were older than 65 years of age, compared to 56.9% of Spanish citizens who tested positive. Migrants from Latin American had higher positivity rates per 1000 people, compared with the host population and other migrant groups (24).

A US study found that being foreign-born was positively associated with COVID-19 case rate at the county level (data to 28 May; with fully adjusted incidence rate ratio 1.106, 95% CI 1.074-1.139; p<0.01) (23).

In Singapore, labour migrants in crowded dormitories have been disproportionately impacted by COVID-19, with over 95% of confirmed cases (to 19 June) among dormitory-housed migrants; as of 18 Nov, 54,502 (95.7%) of 58,135 all in-country cases of COVID-19 were in migrants residing in dormitories (29). A study in one isolated dormitory of 5977 migrant workers (mean age 33 years) in an accommodation centre of 13,000 migrants, 1264 tested positive for COVID-19 (between 11 to 19 April) (25). Similarly in Saudi Arabia, Ministry of Health Data reported that 75% of all people in-country who had tested positive for COVID-19 were migrants (to 7 May) (30).

Data on migrants in detention facilities and reception centres suggest these are high-risk settings for COVID-19. In the US, across 52 facilities run by the Department of Homeland Security (DHS)’s Immigration and Customs Enforcement (ICE) agency as of May 2020 more than 50% of ICE migrant detainees who had been tested were positive (27). The European Centre for Disease Prevention and Control has also highlighted several examples of COVID-19 outbreaks in migrant reception and detention centres in the European Union/European Economic Area (EU/EEA) and the United Kingdom (UK) in a technical report, including Greece, Germany, Malta, The Netherlands, and Portugal, and concludes that whilst there is no evidence to suggest that SARS-CoV-2 transmission is higher amongst migrants and refugees, overcrowding in reception and detention centres may increase their exposure to the disease (28).

This is in line with national notification data from Germany where 2.5% of notified outbreaks up to 11 August (199 of a total of 7864 were reported in refugee centres comprising 7.5% (n=4,146) of all notified cases during outbreaks (n=55,141) across the country. The average number of cases per outbreak in refugee centres was 20.8, higher than in any other outbreak setting (31). A systematic analysis of outbreak reports to 22 May identified 42 outbreaks in refugee reception and district accommodation centres of 11 federal states, with 1781 confirmed SARS-CoV2 cases among 9785 refugees in those centres. The pooled cumulative incidence rate (attack rate) was reported as 17.0% (95% CI 12.0 – 23.0, I_2_ = 98.3%) (32).

A temporal and spatiotemporal dynamics study of the COVID-19 pandemic in Kuwait using daily confirmed case data collected between the 23 February and 7 May concluded that densely populated areas and poor living conditions of migrant workers resulted in the highest number of significant spreading and clustering events within their communities (46).

We found one Italian study reporting no differences between migrants and non-migrants in terms of the probability of being tested (OR 0.93; 95% CI 0.81-1.1) and a similar prevalence of infection (OR 0.99; 95% CI 0.82-1.20) (22).

### Hospitalisation due to COVID-19

In a prospective COVID-19 registry study (n=1123) comparing Kuwaitis with non-Kuwaitis/migrants (two-thirds of the Kuwaiti population are migrants, the majority of non-Kuwaitis are migrant workers) in the main COVID-19-specific healthcare facility in the country, with adjustments made to age, gender, smoking and selected co-morbidities, non-Kuwaitis (91.6% males; mean age 41.0 years) had two-fold increase in the odds of death or being admitted to the intensive care unit compared to Kuwaitis (OR 2.14, 95% CI 1.12–4.32). Non-Kuwaitis also had higher odds of acute respiratory distress syndrome [ARDS] (OR 2.44, 95% CI 1.23–5.09) and pneumonia (OR 2.24, 95% CI 1.27–4.12) (34).

In Denmark, non-Western migrants and their children accounted for 15% of COVID-19 hospital admissions (to 7 September), despite only making up 9% of the population (20).

In one province in Italy (27 February to 2 April), migrants were found to have a higher risk of hospitalisation (hazard ratio [HR] 1.3, 95% CI 0.99-1.81) than Italians (33). In Italian surveillance data (to 19 July) non-Italian cases were diagnosed at a later date than Italian cases and were more likely to be hospitalised (adjusted relative risk 1.39 [95% CI 1.33-1.44]) and admitted to an intensive care unit (1.19 [95% CI 1.07-1.32]), especially in those coming from lower human development index countries (35).

In Greece, almost half of COVID-19 patients hospitalised in Attica (Athens and surrounding areas) as of 17 Sept were refugees from camps/hosting sites and destitute migrants from the city centre, including in Sotiria hospital (40 of 103 are refugees), Evaggelismos (36 of 66), Amalia Fleming (10 of 20) and Attikon (26 of 26); many of these patients were reported to be “asymptomatic and young” but could not be returned to overcrowded accommodation (47).

### COVID-19 mortality and excess deaths

An analysis of all recorded COVID-19 deaths (to 7 May) in Sweden found that being an migrant from an LMIC is predictive of a higher risk of death from COVID-19, but not for all other causes of death (42). In models adjusting for age and sociodemographics, migrants from LMICs from the Middle East and North Africa found a three times higher mortality rate from COVID-19 among men (HR 3.13, 95% CI 2.51-3.90) and two times higher mortality among women (HR 2.09, 95% CI 1.52-2.89) as compared to the Swedish-born (42). Similarly, data from Stockholm, Sweden until 4 May shows that migrants from Middle Eastern countries (RR 3.2, 95% CI 2.6-3.8), Africa (RR 3.0, 95% CI 2.2-4.3) and the Nordic countries (RR 1.5, 95% CI 1.2-1.8) had higher COVID-19 mortality when compared to Swedish-born people, adjusting for age, sex and sociodemographic characteristics. Especially high mortality risks from COVID-19 were found among individuals born in Somalia (RR 8.9, 95% CI 5.6-14.0), Lebanon (RR 5.9, 95% CI 3.4-10.3) and Syria (RR 4.7, 95% CI 3.3-6.6) (43).

An epidemiological report that compared risk of death from COVID-19 in over 25-year olds who were foreign-born versus Swedish-born of the same age to 30 June in Stockholm Country found marked differences between Swedish-born and Somali (HR adjusted for age and sex 12.39 [7.93-19.36]), Lebanese (6.19 [3.41-11.24]), and Syrian (6.14 [4.28-8.80]) migrants (40). These effects were attenuated when adjusted for neighbourhood, education level, occupation, income, household size and previous chronic illness, but remained higher among migrants than Swedish-born (44). In a brief report of 106 healthcare workers who died in the UK up until 22 April 2020, 56 (53%) were reportedly born outside the UK (45).

No differences in mortality from COVID-19 by migration status were observed in crude analyses by migrant status in Denmark (data to 7 September) (20). In one province of Italy, migrants were found to have a similar risk of death to non-migrants (27 February to 2 April) (33). However, Italian surveillance data from the start of the outbreak to 19 July found an increased risk of death in non-Italians from low-Human Development Index countries (adjusted RR 1.32, 95% CI 1.01-1.75) (35).

Definitional and data collection challenges mean that attention has focused on all cause excess mortality during the pandemic, comparing deaths with those expected on the basis of rates in preceding years. In England, for example (21 March to 8 May) where the number of death registrations from all causes was 1.7 times higher than the average during the same period in 2014-2018, the relative increase in total deaths was greater among those born outside the UK; deaths in 2020 were over 3 times higher than the equivalent period in 2014 to 2018 for those from Central and Western Africa (4.5 times higher) the Caribbean (3.5), South East Asia (3.4), Middle East (3.2) and South and Eastern Africa (3.1). For migrants born in other countries within the EU (internal migrants) the level of increased risk was similar to those born in the UK (3).

In France, foreign-born people represented 15% of registered deaths (March and April 2020) versus 13% for the same period in 2019. This includes an increase of 54% deaths among migrants from North Africa (Algeria, Morocco, Tunisia), 114% for those from sub-Saharan Africa, and 91% for those from Asia. Migrants from other parts of Europe, America or Oceania had similar mortality rates to the French-born, who experienced a 22% excess mortality (37). This same trend is also seen in different regions of France; for example Seine-Saint-Denis, a district in the north of Paris where 30% of the population are immigrants, saw a 188% mortality increase compared with 2019, versus a 96% increase in Paris as a whole (38).

In the Netherlands (9 March to 19 April 2020), mortality was 47% higher than expected for migrants from non-Western countries and their immediate children (based on number of deaths in the preceding weeks, adjusted for seasonal factors), 49% higher for migrants from Western countries and their children, and 38% higher for the native-born people with Dutch parents (39).

In Sweden, mortality among migrants was elevated in 2020 compared with previous years. A comparison between all-cause mortality data from March to May 2020 with data from the same period in 2016 to 2019 found that among middle-aged (40-64 years) and older (>65 years) migrants born in Syria, Iraq and Somalia excess mortality was approximately 220%. Among people born in Sweden, the EU, the Nordic countries or North America, the excess mortality among those >65 was 19% and among the middle aged was 1% (40). In Stockholm during the peak of the epidemic (6 to 12 April 2020), areas with the lowest tercile of share of Swedish-born had 178% excess mortality compared with the previous five years (41).

In Italy, on the other hand, between 21 February and 29 April 2020, found the share of migrants and non-migrants among COVID-related deaths (2.5% and 97.5% respectively) was similar to their share in all-cause mortality rates estimated in Italy in 2018 (2.6% and 97.4% respectively) (36). However, migrants were younger at the time of death than non-migrants (71.1, standard deviation [SD] 13.1 years vs 78.3, SD 10.8 years, p<0.001).

#### Indirect health and social impacts

The mental health impact of the COVID-19 pandemic and associated restrictions has been well-documented. Migrants may be particularly affected due to pre-existing risk factors (48, 49) and potential exclusion and social isolation (50), and worsening of pre-existing mental health conditions (51, 52); providing remote therapy for these individuals can be challenging (53). In one Canadian study, however, immigrants were found to be less likely to increase negative health behaviours than Canada-born adults (54). In a nationally representative US survey carried out in March 2020, COVID-19-related fear and associated anxiety and depressive symptoms were higher for migrants compared with the US-born (p<0.001) (55), with similar findings in other studies (56, 57). In a cross-sectional survey of 295 Filipino domestic helpers in Hong Kong, multivariate regression results showed that the insufficiency of personal protective equipment (PPE) (OR=1.58 [95% CI 1.18-2.11]), increased workload (OR 1.51 [95% CI 10.2-2.25]), and concerns about being forced out of their jobs if they test positive for COVID-19 (OR 1.32 [95% CI 1.04-1.68]) were significantly associated with anxiety in a multivariate analysis (58).

Migrants may be especially impacted by travel restrictions (8, 9, 54). Arriving migrants have been pushed back or quarantined at borders and forced to stay in informal or overcrowded transit sites, while international refugee resettlement programmes have been disrupted (8, 59). For migrants who are already settled, but not considered resident, border restrictions may force them to overstay their visas, or prevent them from visiting family or friends outside of their host country, exacerbating feelings of isolation (60). Concerns have also been raised that border closures may increase smuggling of migrants (61). COVID-19 may meanwhile pose a barrier to integration for migrants and refugees (62), for example due to the suspension and modification of resettlement schemes (63, 64), and education programmes (63, 65–68). Migrants who are particularly vulnerable may be disproportionality affected by the negative social impact of lockdown (69, 70). Migrants are considered to be especially vulnerable to job loss and economic hardship as a result of COVID-19 (60, 63, 67, 71–77). A qualitative cumulative risk assessment for migrant workers in Kuwait found many workers are now facing layoffs, furloughs, non-payment and late payment of wages putting them in significant financial hardship (78). Across Organisation for Economic Cooperation and Development (OECD) countries, approximately 30% of migrants are considered to be living in relative poverty, compared with 20% of the native-born people (67), which increases their vulnerability to COVID-19 infection (50, 79).

Migrants may also be experiencing discrimination as a result of the COVID-19 pandemic (8, 80, 81). In particular, Chinese and other Asian migrants have been targeted due to the original emergence of the pandemic in China, with reports of bullying, awkward behaviour, avoidance of Chinese restaurants and shops, and physical attacks (82–84). In surveys and interviews with people of Chinese origin living in France, nearly a third reported having experienced at least one discriminatory act since January 2020 (85).

#### Risk factors and vulnerabilities for COVID-19 in migrants

Table 2 summarises key risk factors for migrants for COVID-19 reported from included data sources. Figure 2 highlights key risk factors and vulnerabilities of migrants identified in the literature.

**Figure 2:**
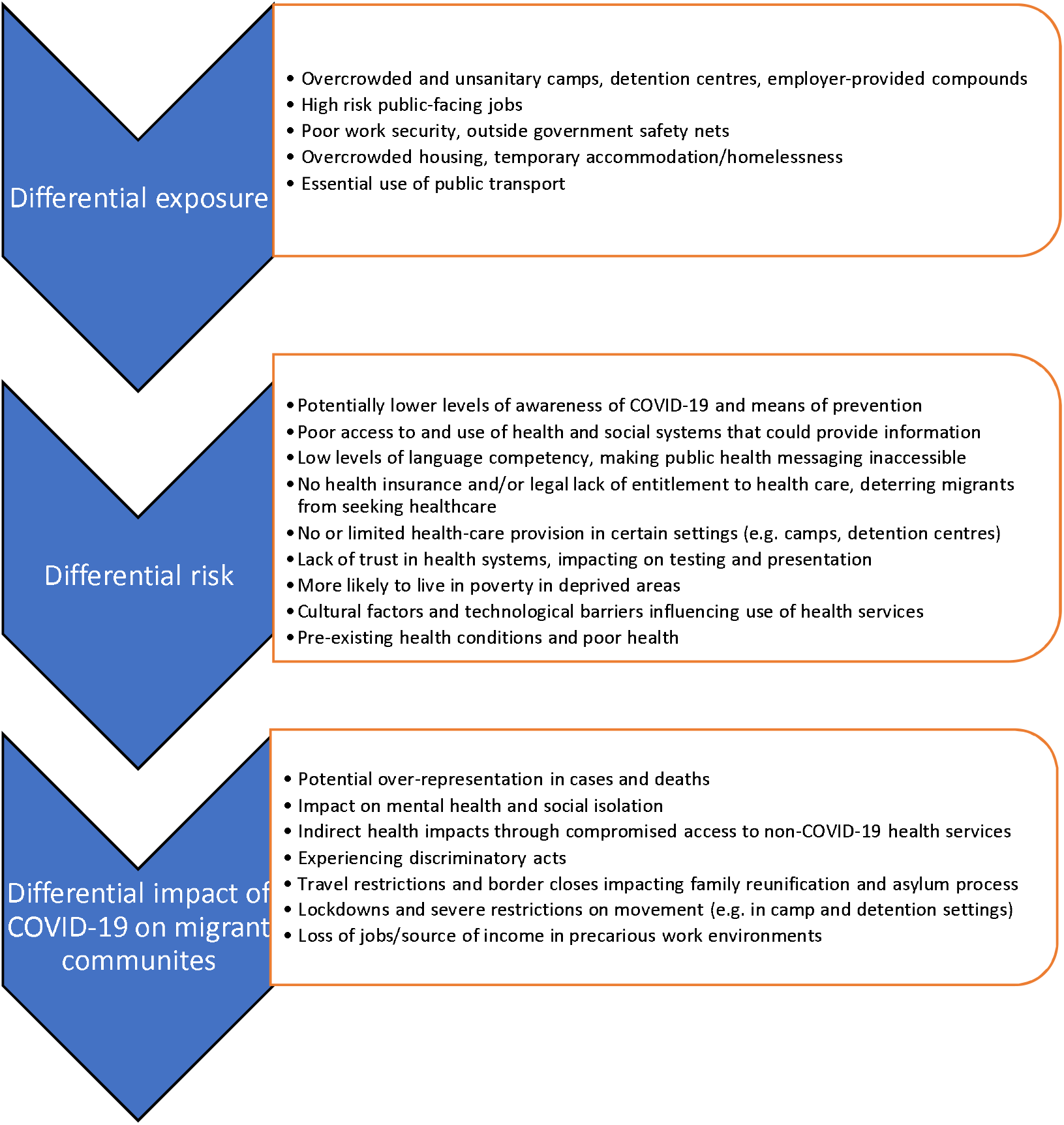
Migrant-specific risk factors and vulnerabilities identified in included literature.

**Table 2:**
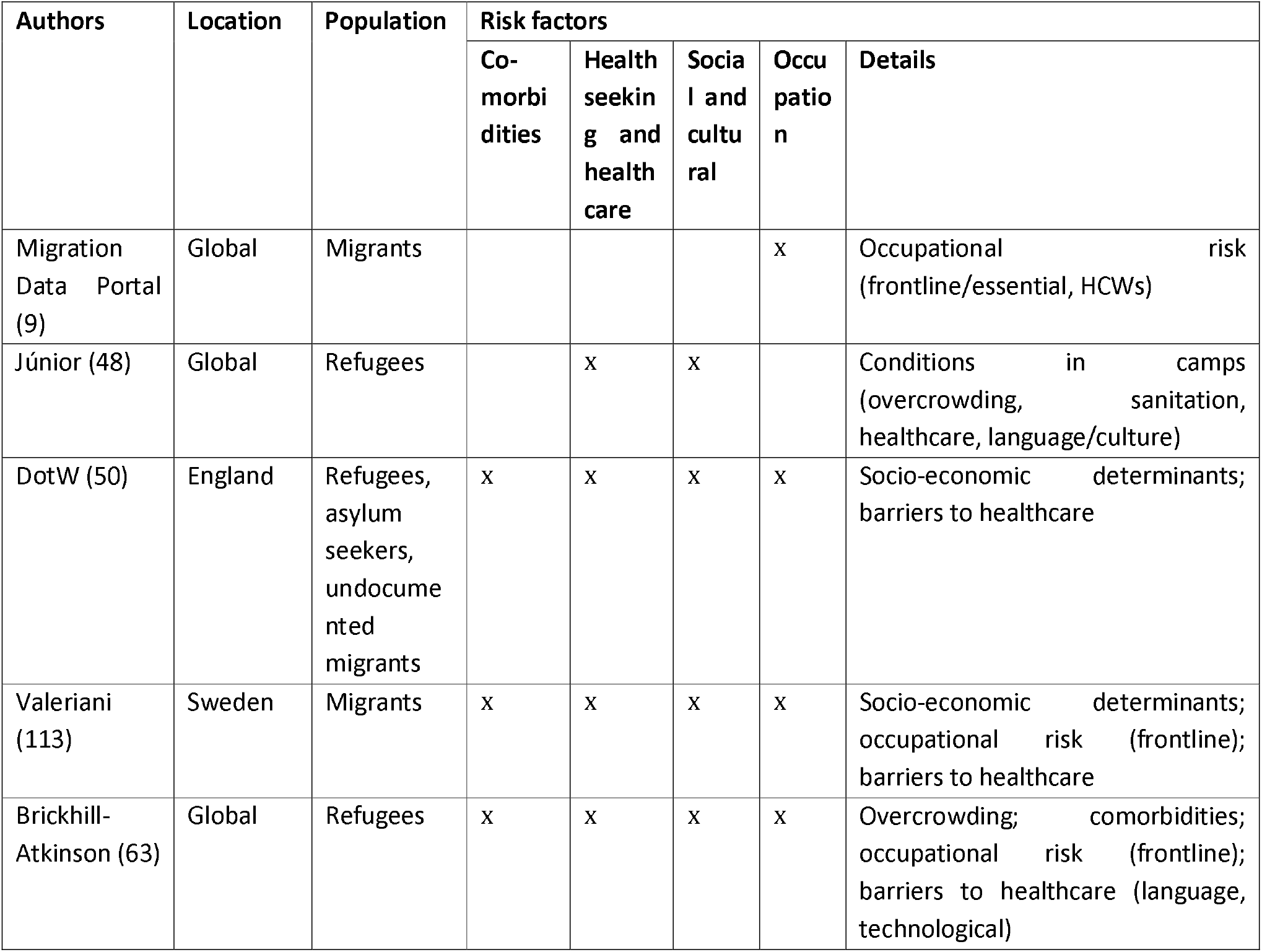
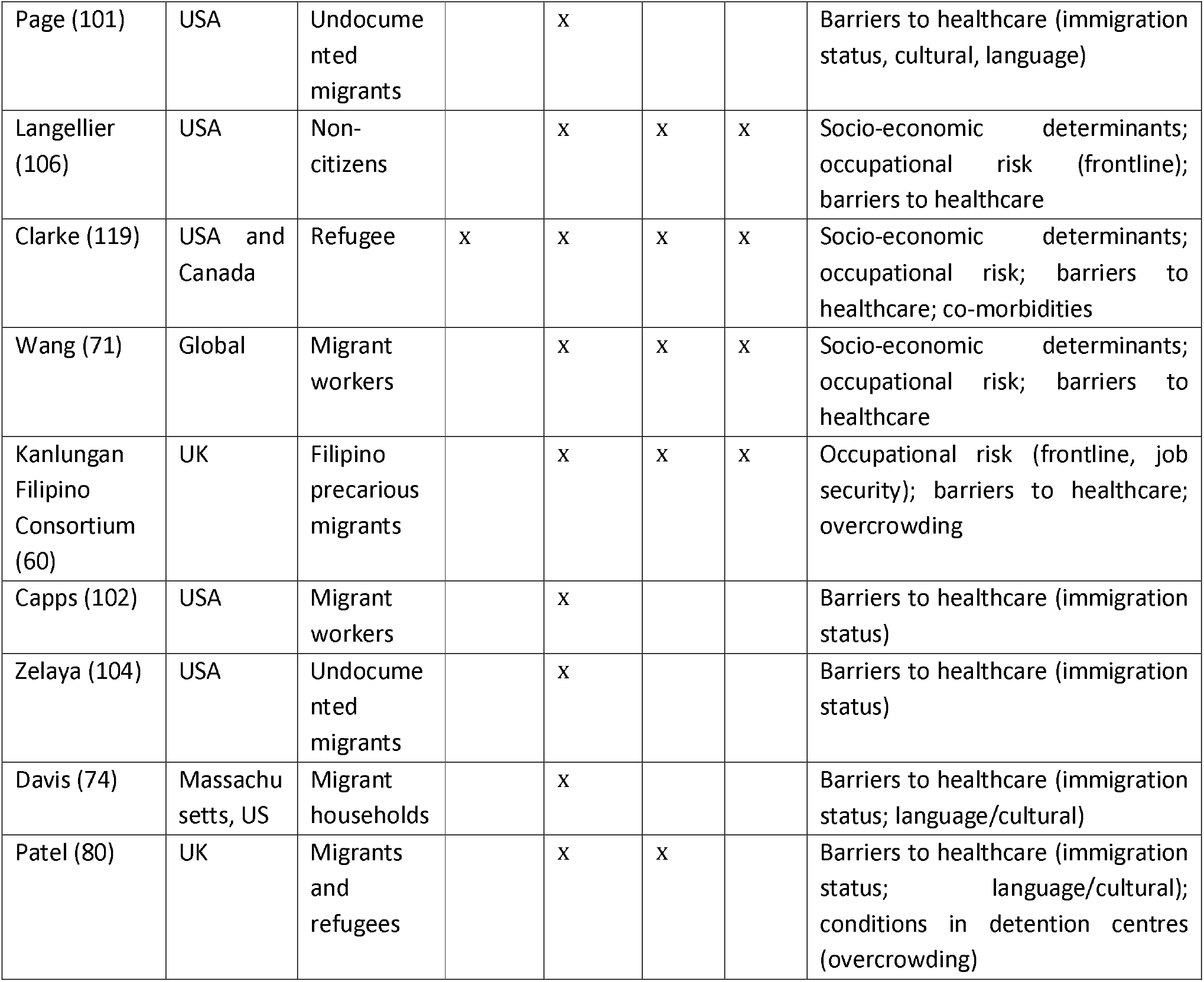
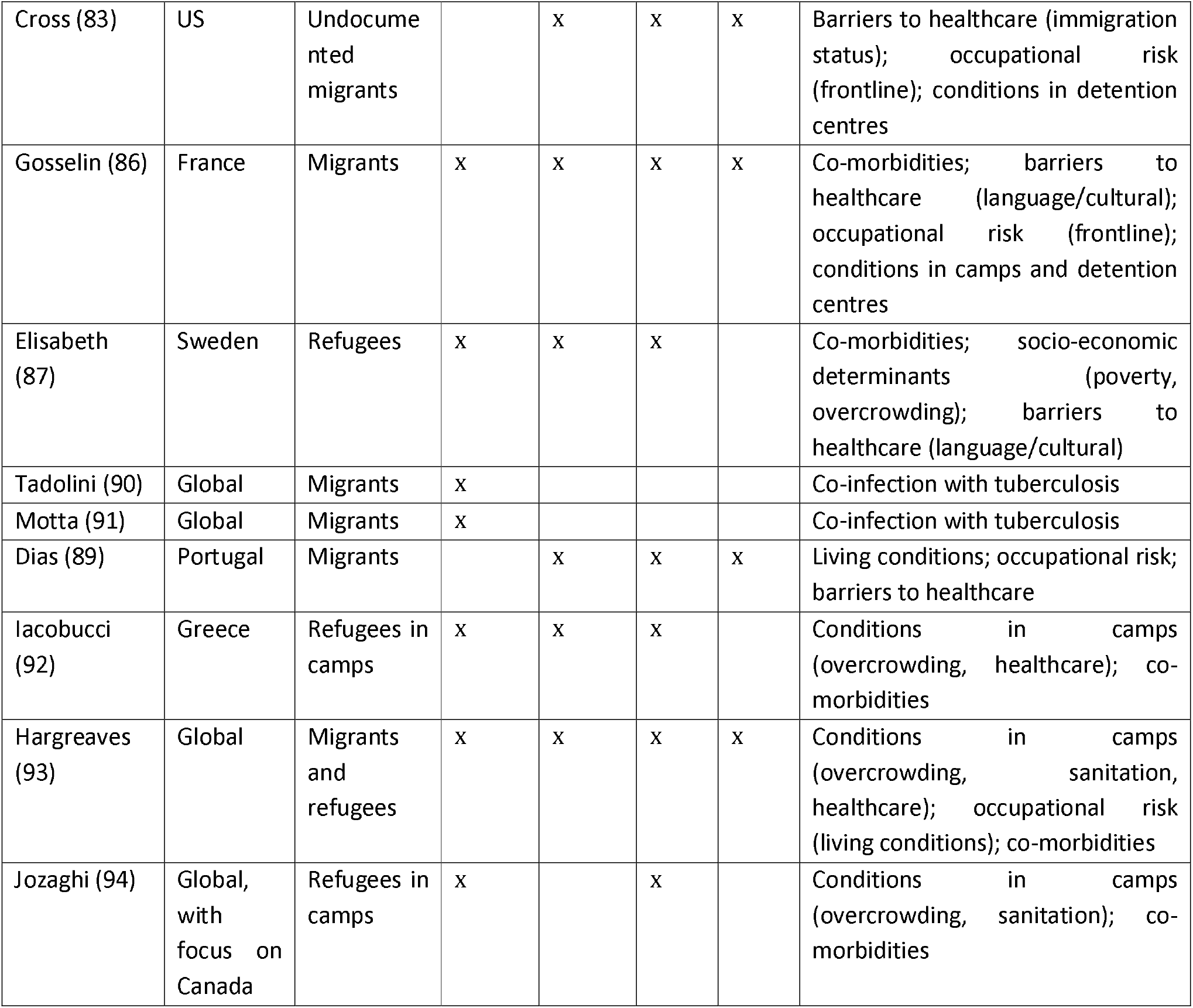
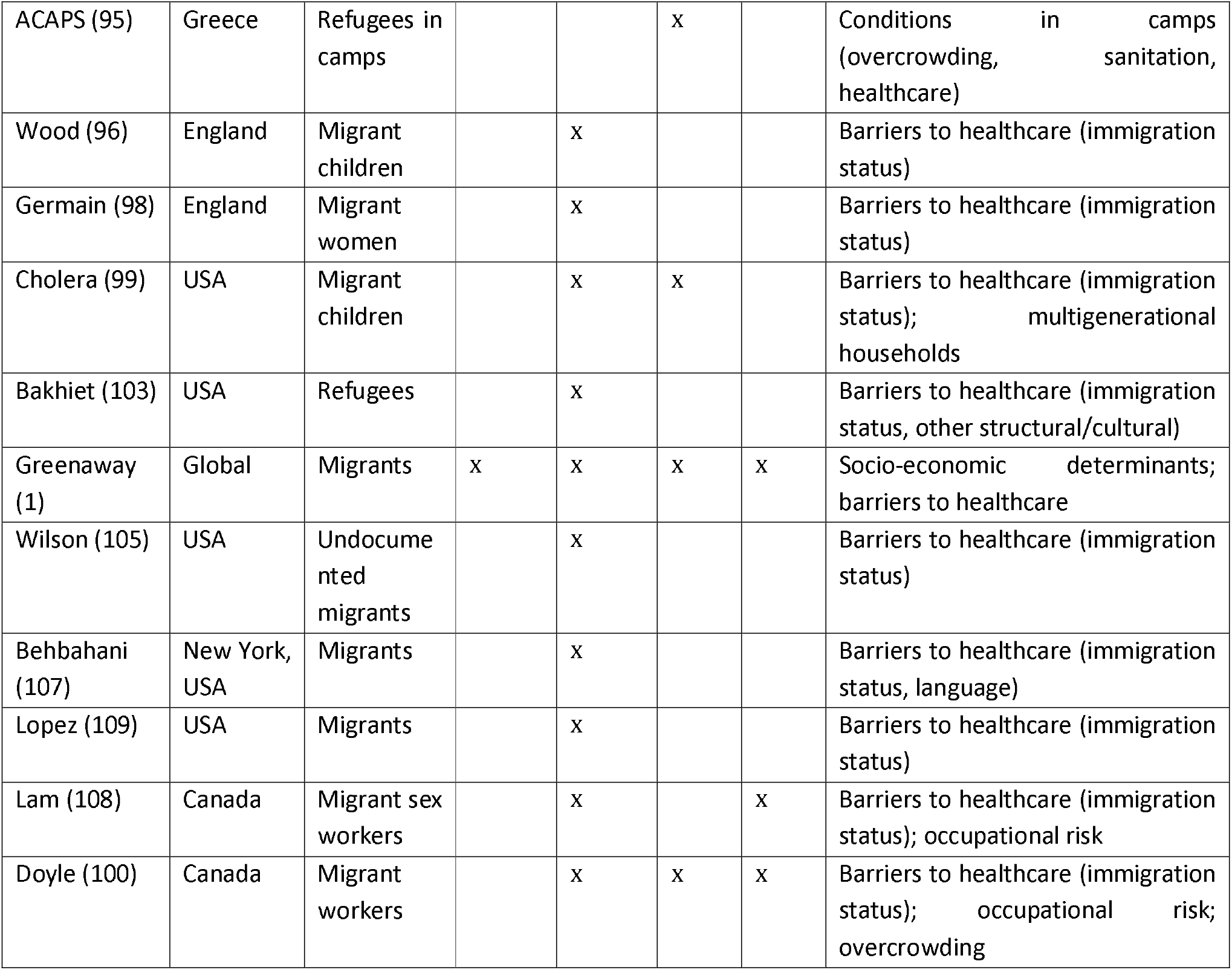
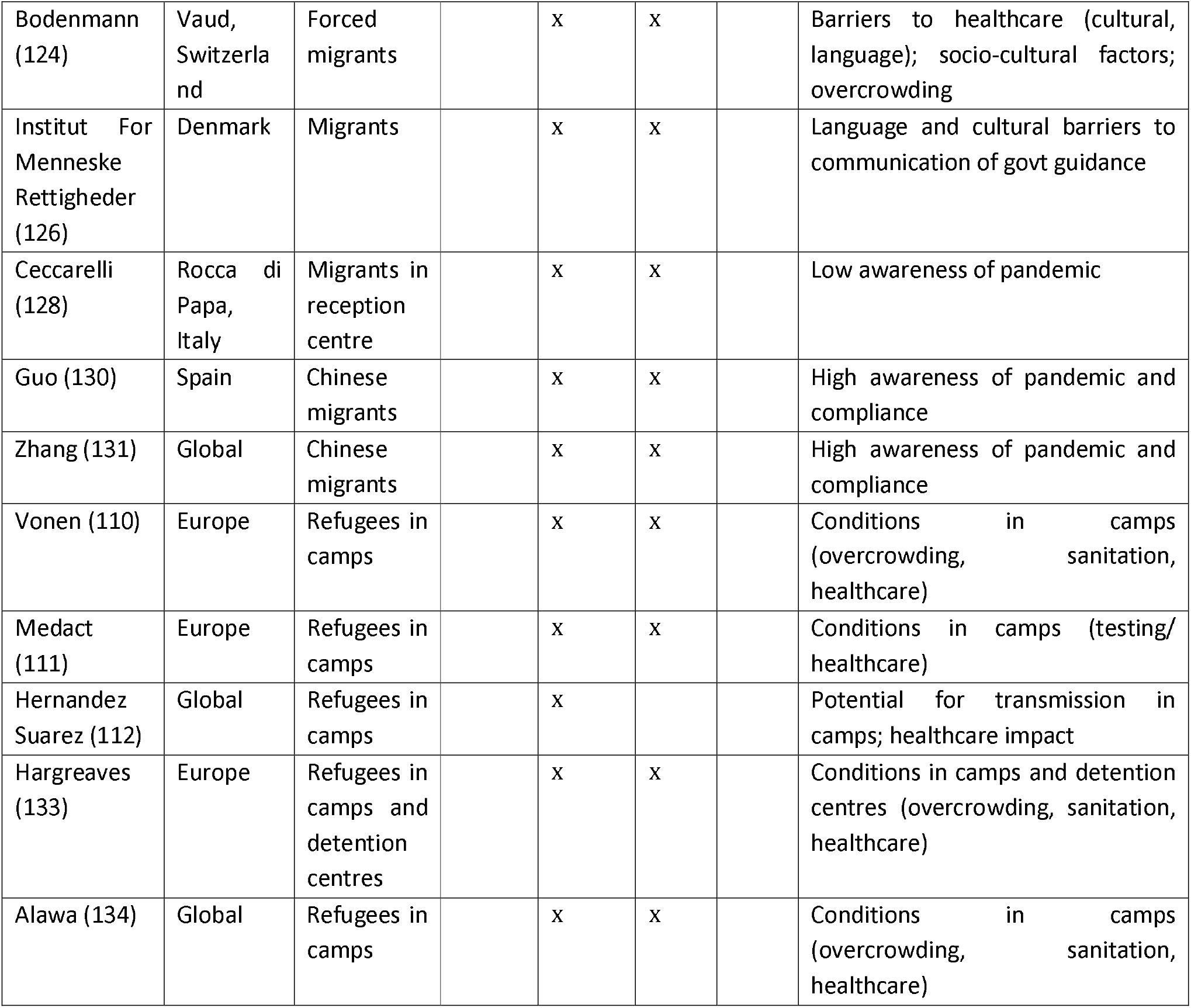
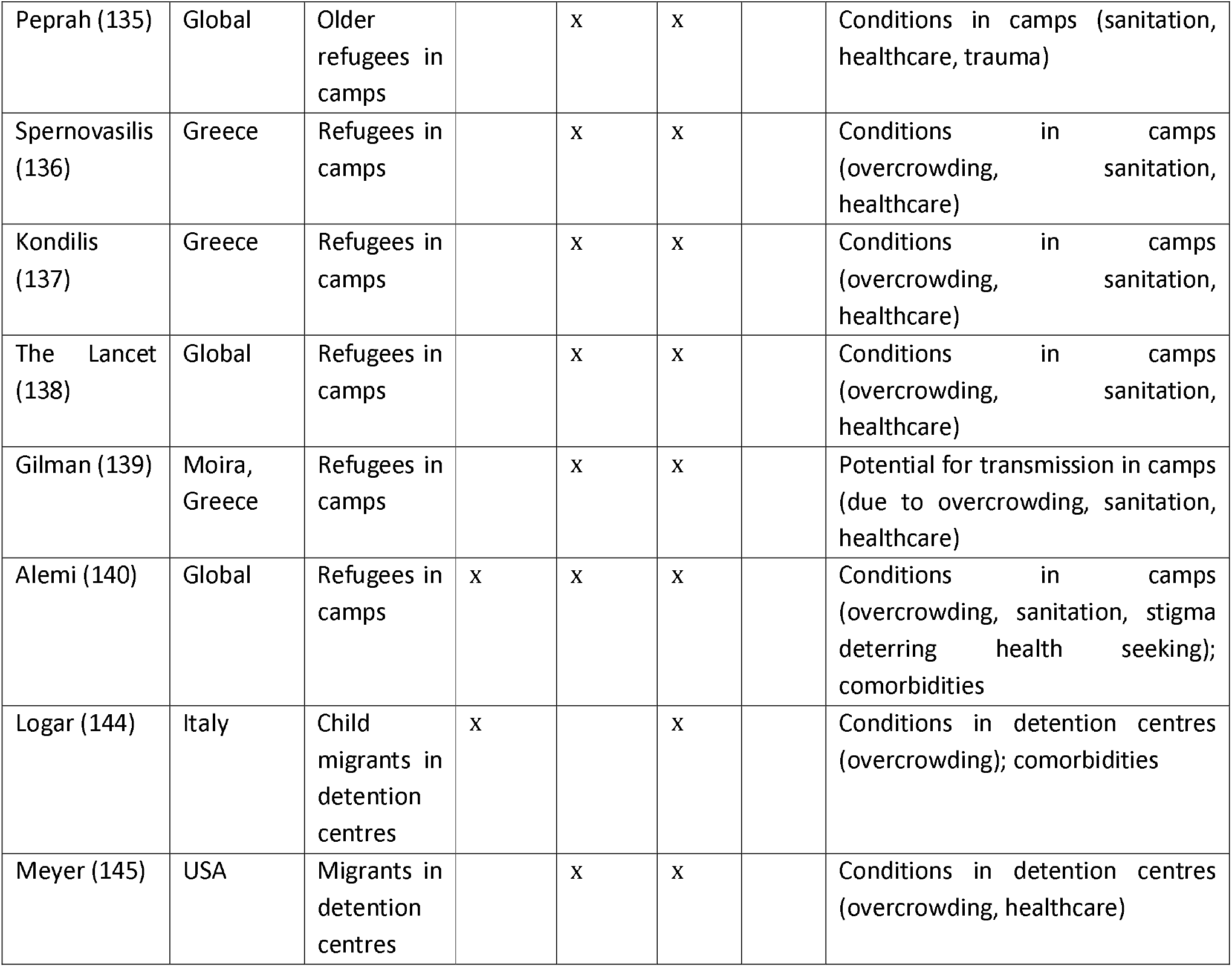
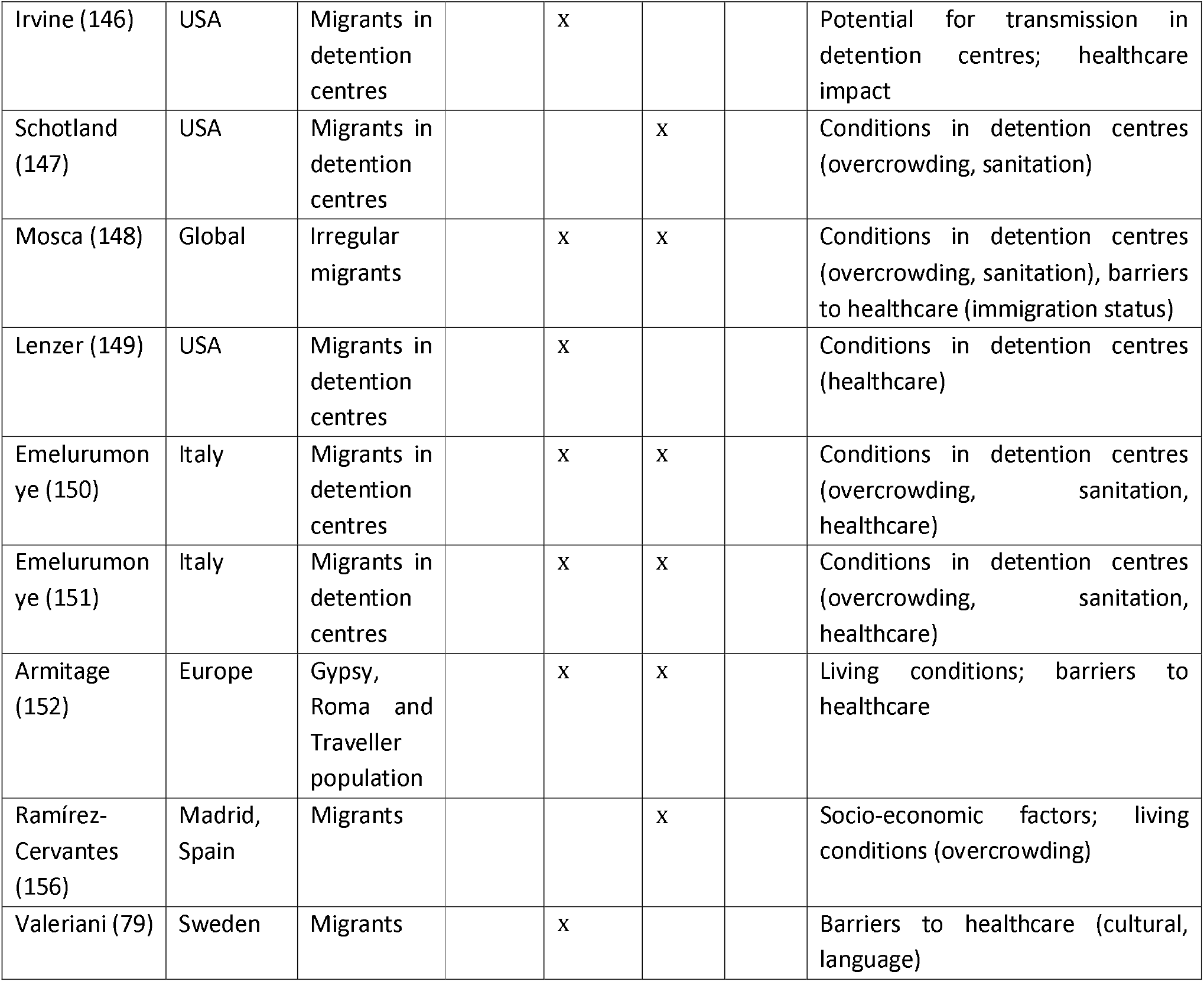
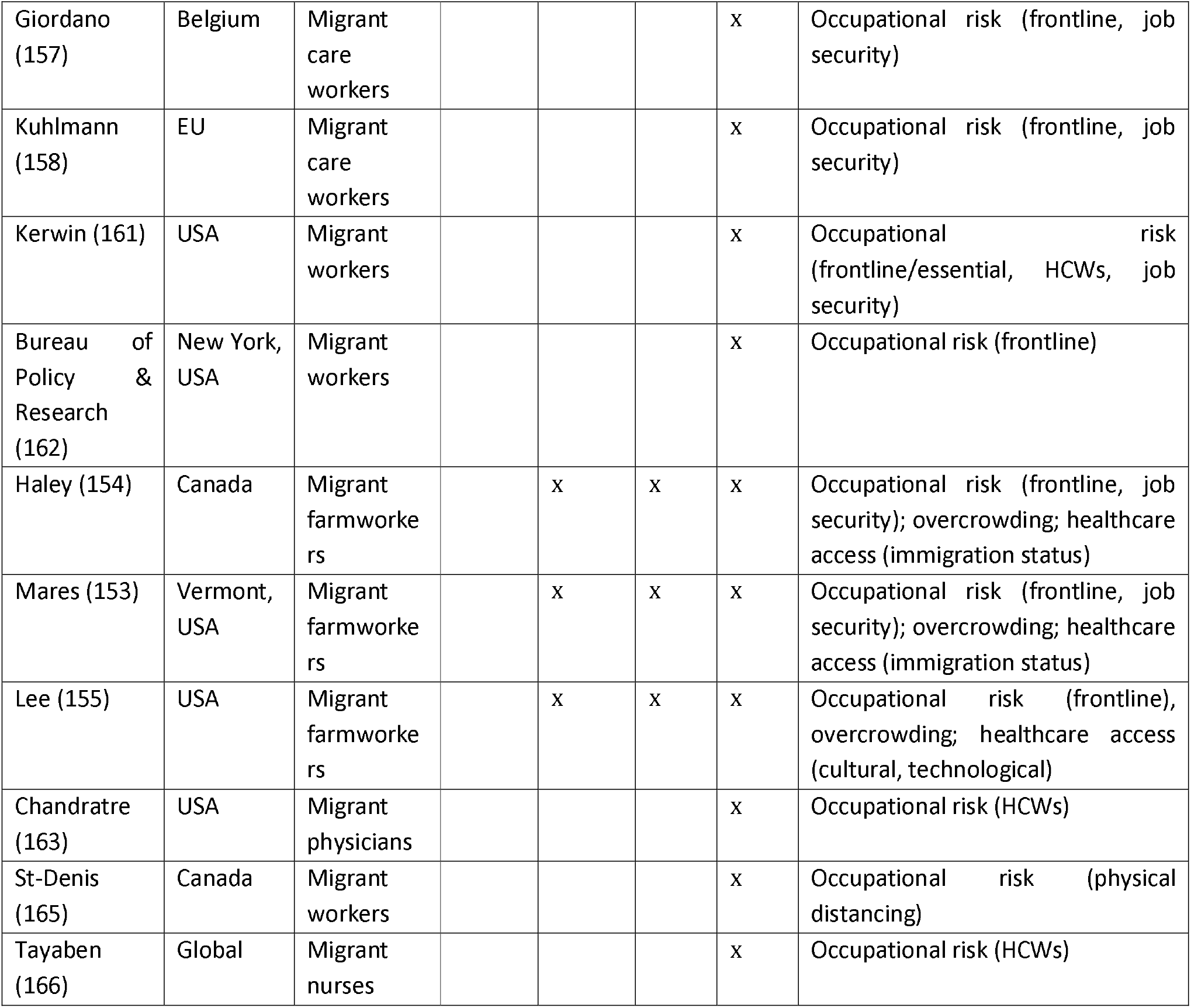
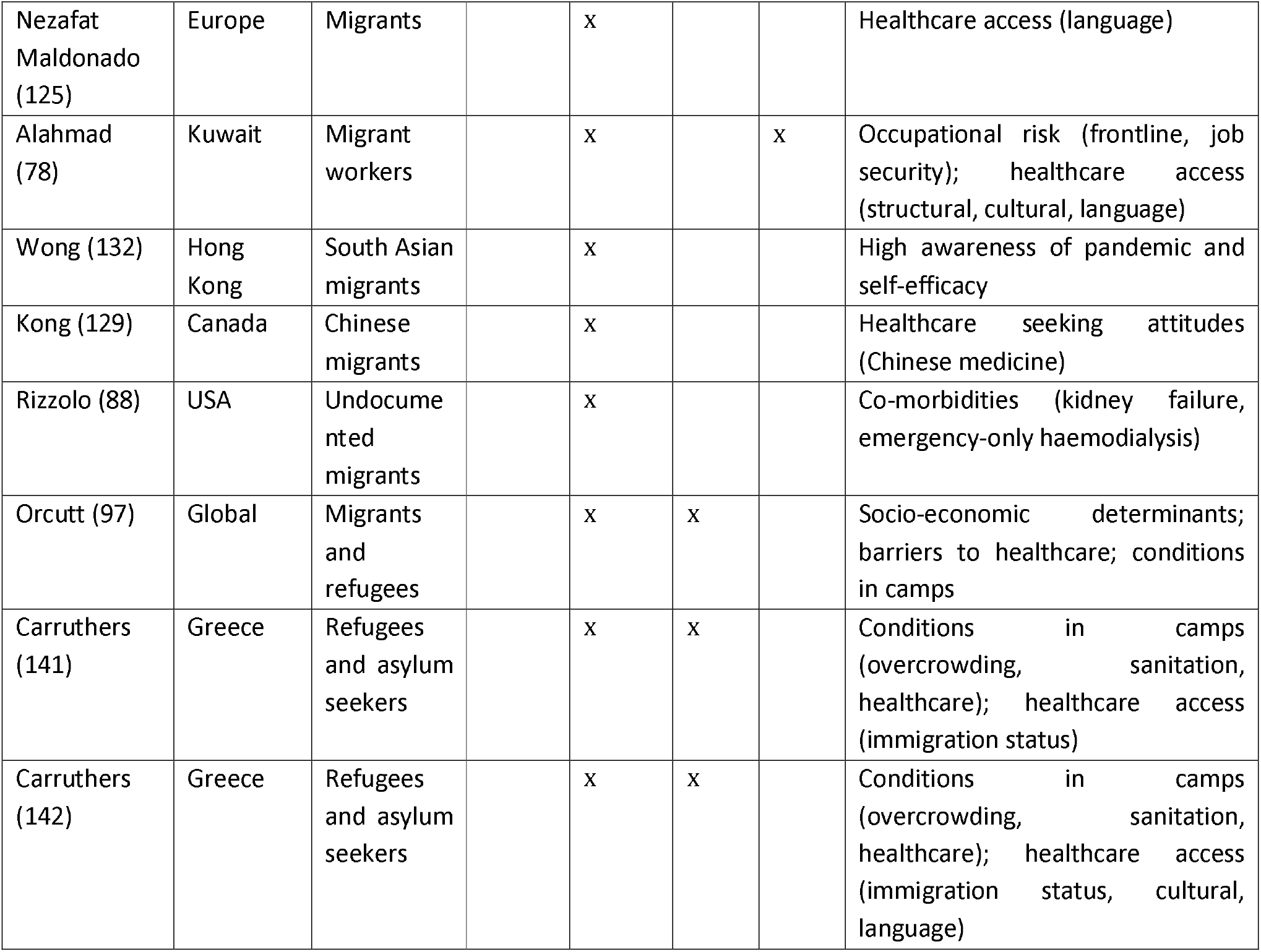
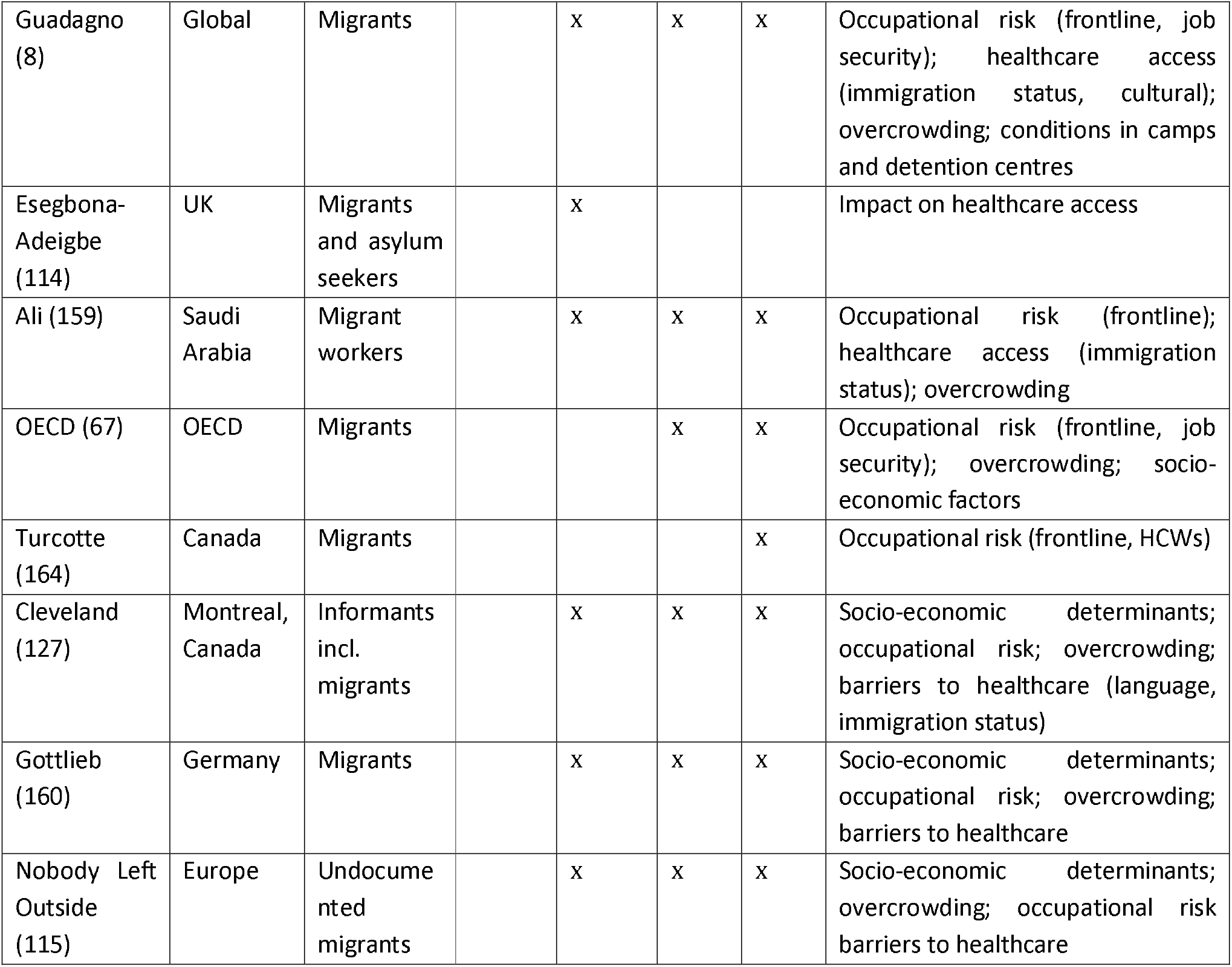
Risk factors and vulnerabilities reported for migrants for COVID-19.

### Co-morbidities

Co-morbidities may be a cause of increased COVID-19 risk and/or poor COVID-19 outcomes in migrant populations, but this remains poorly documented. A situational brief reporting on the health or asylum seekers and undocumented migrants in France during COVID-19 concludes they are more likely to have certain chronic conditions that appear to be associated with worse COVID-19 outcomes, such as diabetes mellitus, hypertension, and obesity (86). In Sweden, a COVID-19 situational report found around 65% of refugees are either overweight or obese compared to 50% in the rest of the population, and around 35% are smokers, which is higher than the general population (87). In addition, hospital visits for management of co-morbidities may increase risk of exposure to COVID-19 (88). Co-infections may also play a role. In Lisbon, it has been observed that some of the neighbourhoods with increased transmission coincide with areas where TB incidence has been higher (89), with over half of patients with TB and COVID-19 in two early case series being migrants (90, 91). Migrants in camp settings may be especially vulnerable due to existing illnesses or injuries and prevailing malnutrition and/or poor health in general (92–95).

### Healthcare seeking and barriers to care

Testing and treatment for COVID-19 has been made free of charge and exempt from immigration status checks in many countries, with these messages communicated in multiple languages; however, concerns remain that these exemptions do not fully mitigate the extensive barriers that migrants experience in accessing healthcare (8, 96, 97). Concerns within migrant communities that COVID-19 treatment might be chargeable, or that undocumented migrants might be identified by health systems on presentation remain, and could prevent early presentation and testing in migrants who distrust authorities (50, 60, 98). In the US, where nearly half of undocumented adult migrants and a quarter of lawfully present adult migrants lack health insurance (99), or have insurance that relies on a specific employer, migrants may avoid seeking care for fear of losing their job and being deported (100). Various federal policies deter migrants from health seeking (101–104). For example, undocumented migrants in the US are ineligible for federally funded healthcare programmes such as Medicare and Medicaid (105), and the ‘public charge’ rule introduced in February 2020 makes migrants who receive a broad range of cash and noncash benefits ineligible to apply for citizenship and residency (106, 107), deterring treatment-seeking, particularly so in jobs that are often criminalised such as sex work (108). US Immigration and Customs Enforcement (ICE) raids have continued in migrant communities over lockdown, and have further damaged trust and deterred migrants from testing and treatment (109).

In an online survey of undocumented migrants (students who entered as minors) in the US (May 2020), 10% said that they or an immediate family member suspected COVID-19 infection at some point but did not get tested for fear of detainment or deportation, and 1 in 5 said they would be ‘extremely worried’ for this reason (56).

Healthcare access for migrants and refugees in camp settings can be limited, lacking medical personnel, equipment and pharmaceuticals (110), with poor or absent testing facilities (111). A modelling study has suggested that once the virus enters refugee camps, it can spread quickly, overwhelming hospitals and healthcare facilities (112).

With routine services closed due to the pandemic, concerns have been raised that migrants have struggled to navigate the new systems (50, 113–115) and it has exacerbated migrants’ exclusion from health services (116–120). Migrants may experience challenges in accessing healthcare remotely (50, 63, 101, 106, 121); however, telemedicine may also offer opportunities that need to be further explored in these populations (122, 123).

Migrants often have difficulties understanding public health messaging due to cultural and language barriers (50, 99, 124). Public health guidance in many countries was not initially tailored to the needs of migrant and ethnic minority groups (50, 125); in the UK non-governmental organisations (NGOs) translated material into 51 languages to make it more accessible (80). In Denmark, a series of qualitative interviews with migrants found that they felt uncertain regarding government guidance for COVID-19; although written material was translated into 19 languages, it was not effectively disseminated (126). In Montreal, Canada, it took two months after lockdown started for the Public Health directorate to publish official multilingual fact sheets on COVID-19 guidelines, and information phone lines only operate in French and English. Those who had arrived most recently, had lower language (French/English) ability or lower literacy had more difficulty accessing local COVID-19 information (127). In a rapid review to assess communications targeting migrant populations across Council of Europe Member States only 48% (23/47) translated information into at least one foreign language (125). Information on testing or healthcare entitlements in common migrant languages was only found in 6% (3/47) of countries and no government produced risk communications on disease prevention targeting people in refugee camps or informal settlements. Poor language competence linked to low testing rates in two studies (13, 15).

A potential lack of knowledge and awareness of COVID-19 among migrant groups or spread of misinformation has been reported (50). For example, in qualitative interviews conducted in a migrant reception centre in Rocca di Papa, Italy between February and July 2020, there was low awareness of the danger of the pandemic, especially among migrants from sub-Saharan Africa (128). There is some evidence that traditional Chinese medicine may have been used as a means of preventing COVID-19 among Chinese immigrants in Canada (124, 129). Conversely, migrants may be more likely to comply with preventative measures such as mask wearing, especially those migrating from Asian countries where this is more of a cultural norm (130, 131). A questionnaire among 352 Indian, Pakistani, and Nepalese migrants in Hong Kong found migrants expressed certain misconceptions regarding the prevention of COVID-19 infection, but perceived the risk of disease as mild, had positive attitudes regarding its prevention, and implemented recommended disease-preventive measures (132).

### Camps, detention centres, and overcrowded accommodation

Refugee camps are typically crowded, and are often built quickly and with little regard to such things as tent spacing. In these settings, where social distancing and personal hygiene is difficult, the spread of COVID-19 is facilitated (1, 92–95, 110, 133–143). For example, the Moria camp in Greece had an estimated population density of 133,000 per km^2^, with reports of one water tap shared between 1,300 people in some areas of the camp (95).

Conditions in detention or reception facilities are similarly conducive to the spread of COVID-19, with confined and poorly ventilated spaces (50, 83, 144–148). In the US, there have been concerns that ICE facilities have violated their own standards as well as those from the Center for Disease Prevention and Control (CDC), for failing to test sick detainees (27, 149). Living conditions in reception facilities in Europe are overcrowded (28, 150, 151). Gypsy, Roma and Traveller populations are also at risk due to living in potentially crowded conditions, their nomadic way of life, and reduced engagement with health services (50, 152). Many migrant workers live in employer-provided shared accommodation, considered high-risk for COVID-19 (100, 153–155).

Migrants in the community are more likely to live in shared or overcrowded accommodation than non-migrants in host countries (50). 235 (59%) of 399 of patients admitted to a medicalised hotel in Madrid in March to May 2020 were migrants: the main reason for referral was a lack of housing that supported quarantining, for example due to overcrowding, which was correlated with migrant status (χ^2^ =19.4, p<0.01) (156). At a clinic in Milan, the proportion of undocumented migrants who were homeless nearly doubled from 8.8% to 17.1% in the months immediately before and during/after lockdown (120). In a survey of precarious Filipino migrants in the UK, most of whom were undocumented migrants, 58% of respondents lived in shared houses, 1 in 5 were homeless, had no fixed address, or were staying temporarily with friends (on average sharing a bedroom with 1-2 others) (60).

Across all OECD countries, migrants are more likely to live in sub-standard accommodation (23% versus 19% in the native-born) and twice as likely to live in overcrowded dwellings (17% versus 8%) which could influence transmission and exposure (67). Living in neighbourhoods with higher household density was associated with higher positivity rates for COVID-19 in Ontario, Canada, but especially for migrants (13). Migrants are also more likely to live in multigenerational houses, with implications for transmission from younger to older and more vulnerable household members (99, 106).

### Occupational risk

Migrants are disproportionately represented in front-line public-facing jobs, such as in the fields of healthcare, social work, hospitality, retail, delivery and household services, and in menial jobs that can place them at increased exposure of COVID-19 (8, 83, 154, 157–160). On average, 13% of all key workers in the EU are immigrants (9). Based on 2018 US Census Bureau data for a report on COVID-19 impacts, 69% of all immigrants in the US labour force and 74% of undocumented workers were reported to be essential workers, compared to 65% of the native-born labour force; 70% of refugees and 78% of Black refugees are essential workers (161), with non-US-citizens making up 9% of the labour force but 22% of workers in the agricultural industry, for example (106). In New York, the hardest hit US city during the first wave of the pandemic, 50% of non-governmental frontline workers are migrants (162).

Migrants may need to carry on working or risk losing their job (60, 157). This is especially true for migrants in informal ‘no work, no pay’, with precarious contracts, or exploitative employment, including undocumented migrants who fall outside of government safety nets (60, 162). A Canadian analysis found that workers in low-income occupations (especially women, migrants, and visible minority groups) are employed in occupations that put them at greater risk of exposure to COVID-19 than other workers; low-income workers may face financial disincentives for absence even if they are sick or vulnerable, increasing workplace transmission (152). Migrants are also potentially more likely to rely on public transport to get to work, again increasing their possible exposure to COVID-19 infection (106).

Not all migrants are unskilled or work in low-skilled occupations, however. A substantial proportion of doctors, nurses, and other medical specialists in countries such as Germany, France, US, Canada, and UK are migrants (9, 163). In Canada (2016 data), more than a third of nurse aides, orderlies and patient service associates were migrants, with Black and Filipino women particularly over-represented (164). Data are lacking on the impact of COVID-19 on this occupational group, and on hospital cleaning and maintenance staff who in many EU countries also tend to be migrants. In a Canadian analysis, migrants in health occupations were found to have a slightly higher mean occupational risk of exposure to diseases/infections such as COVID-19 than Canadian-born workers (165). Employment as a healthcare worker in Ontario accounted for a disproportionate number of COVID-19 cases among migrants, especially women (13). Concerns have also been raised about inadequate access to or use of PPE, overrepresentation of migrants in low paying paramedical roles, or difficulties in self-isolating because of staff shortages at the start of the pandemic (1, 166).

Living in low-income neighbourhoods was strongly correlated with test positivity for newly-arrived migrants but not for Canadian-born and long-term residents (13). In addition, the association between percentage of immigrants living in a given area of Ontario and diagnoses of COVID-19 is attenuated when adjusting for covariates such as household income, educational attainment, and household density (14). In Swedish data, socioeconomic status (including disposable income and employment status), number of working age household members and neighbourhood population density attenuated up to half of the increased COVID-19 mortality risk, but not all-cause mortality (43), indicating that these factors play a role but cannot account entirely for the observed disparity.

## Discussion

This systematic review is the first attempt to bring together global datasets on the impact of COVID-19 on migrants, and to assess the critical risk factors and vulnerabilities involved, in what is a rapidly evolving field. We found that migrants are at high risk of COVID-19 infection and over-represented in confirmed COVID-19 cases, with data suggesting an elevated risk for COVID-19 among undocumented migrants, migrant health and care workers, and migrants housed in camps and labour compounds. Available data point to a similarly disproportionate representation of migrants in reported COVID-19 deaths, as well as increased all-cause mortality in migrants in reporting countries in 2020, though data are limited. In general, migrants were found to have higher levels of many of the risk factors and vulnerabilities for COVID-19, as a result of increased exposure due to high-risk or precarious occupations, overcrowded accommodation, legal-administrative barriers to healthcare services and low levels of language competence, all of which have a potentially negative impact on awareness of the problem and/or ability to take remedial action – including testing uptake and activities to reduce exposure. These data are of immediate relevance to national public health responses, be it in terms of policies or programmatic actions tailored to reach migrants.

In the most recent and largest systematic review of 18,728,893 patients in datasets reporting clinical outcomes for COVID-19 by ethnicity (42 studies from the US, 8 from the UK to 31 Aug), authors report an increased risk of infection in Black and Asian ethnicities (Asian pooled adjusted RR 1.50 [95% CI 1.24-1.93]; Black 2.02 [1.67-2.45]) compared to White individuals, with Asian individuals being at higher risk of hospital admission to intensive care and risk of death, even after adjusting for confounders such as age, sex, and co-morbidities Other research has highlighted high seroprevalence rates for COVID-19 in people living in precarious situations, suggesting over-exposure of marginalised groups (167). These datasets will include migrants as a subpopulation, but do not disaggregate by migrant status. Our analysis suggests that migrants specifically have an increased risk of infection and points to striking increases in all-cause mortality data among certain migrant groups in the few countries that have reported on this in 2020. More robust data on cases, testing uptake, hospitalisations and deaths from COVID-19 among migrants is therefore warranted and considered urgent. There is also a need to strengthen data systems in HICs so as to better understand the distribution of particular health outcomes in migrant populations, not only with respect to COVID-19 but in other disease areas as well.

In this analysis, we report data that defines a unique set of risk factors and vulnerabilities experienced by migrants in HICs that are influencing exposure and outcomes to COVID-19. These risk factors and vulnerabilities are, in large part, related to their health and social situation in the host country, and the barriers to accessing health systems (including preventative testing and treatment) that they face, which have been well reported for other infectious diseases (168). Risk factors include lower levels of language proficiency rendering public health messaging inaccessible. Low host country language competence, which is particularly the case with more recently arrived migrants, was seen to be associated with lower rates of testing in two studies, but higher rates of positivity when tested (13, 15). We know that few countries specifically targeted public health messaging to migrants, which could have resulted in their exclusion from the larger public health response (125). Precarious occupations and social situations mean that public health proposals such as work-from-home, self-isolation, avoidance public transport, and rapid testing uptake are not relevant for many migrants and point to a type and degree of exclusion or restricted access to mainstream health systems.

Tens of thousands of migrants in HICs are excluded or restricted from accessing mainstream health systems because of their immigration status, likely a major barrier to accessing testing and treatment, and eventual vaccine roll out. Previous data for other infections has shown migrants may be late presenters to health services (169), presenting only where necessary due to concerns around immigration and lack of trust, lack of knowledge of the health system, and barriers to registration and access. These findings strongly support arguments for more tailored and targeted public health initiatives to these groups, including information and communication around testing, contact tracing, isolation, and when to present, as well as tackling misinformation. Actions on behalf of migrants should be undertaken with and through trusted community channels, and developed through direct engagement with at-risk migrant groups. Several groups have called for the temporary suspension of policies that exclude migrants from health systems during the pandemic (97), something several countries have done, along with stressing the importance of inclusion of these groups in ongoing protective measures, information campaigns and health services provision (170, 171). WHO and other agencies have reinforced the need to ensure migrants in camps and closed facilities are offered screening, triage testing, and provided with care (163). UN agencies have also stressed the human rights of refugees and migrants and the need to ensure that COVID-19 responses respect these rights (172).

This review has some inevitable limitations. It was not possible to engage an expert in every HIC and as a result some national statistics and grey literature may have been missed. However, we engaged widely through our international networks to source local literature, and the WHO database sources both peer-reviewed, pre-prints, and grey literature from a diverse range of databases that would not normally have been searched individually for a systematic review. We are therefore confident that we have included the majority of datasets available to 18 Nov. In addition, we have included non-peer reviewed grey literature and pre-prints in the narrative synthesis with obvious limitations. Due to the rapidly evolving nature of the pandemic we felt this was justified and strengthens our description of the current situation facing migrants in HICs. In Table 1 and Supplementary Table 1 we have clearly stated all data sources and have given a quality appraisal score to them.

Panel 1 sets out some of the implications of this work for further research and for health policies. Understanding the lived experience of marginalised migrants will be vital to tackling issues around barriers to care (including of migrants with long-term symptoms), testing uptake, and obstacles and facilitators to eventual COVID-19 vaccination and ensuring good vaccine coverage of, and uptake by migrants and ethnic minorities (173). We believe our findings are of immediate relevance to the ongoing public health responses and should inform policies seeking to minimise exposure to COVID-19 in migrants and ensure their inclusion, through innovative and nuanced solutions with community engagement at their centre.

### Panel 1: Further research and next steps

##### Strengthen data collection and future planning

- Initiate large retrospective and prospective studies, disaggregating by migrant status, exploring disparities in testing and diagnosis, hospitalisations, and COVID-19-related deaths in migrants
- Collate and conduct ongoing analysis of data on COVID-19 vaccine uptake by migrants when vaccine roll out starts, to identify disparities early on so they can be addressed
- Ensure more consistent and complete incorporation of migrant status in surveillance and health information systems taking into account gender, ethnic, linguistic, educational and occupational diversity in migrant populations
- Create more empirical evidence on the link between risk factors identified in migrants and the role they play in driving disparities in clinical outcomes
- Development of pandemic preparedness plans that address migration and migrants, and can be shared by countries

##### Delivery of more effective public health messaging

- Co-produce carefully researched messaging on COVID-19 prevention, testing and treatment, contact tracing, and self-isolation with affected communities, tailored to different cultural and social realities and that considers the unique risk factors and vulnerabilities of migrant populations and offers them meaningful solutions and support mechanisms to reduce their exposure
- Ensure rapid quality translation and more effective dissemination of public health messaging and directives into common migrant languages
- Engagement of diverse high-risk migrant communities, through localised support and community champions, in defining how best to deliver credible information and support on COVID-19 testing, reducing their exposure, social support, and facilitating vaccine roll out, alongside exploring mechanisms to build trust in health systems and tackle misinformation

##### Better consider specific migrant groups

- Proactively include extremely marginalised migrants living in camps, reception centres, detention centres, labour compounds, and undocumented migrants and others facing known structural barriers to health systems in the COVID-19 response

##### Long-term approaches to tackling disparities facing migrants in HICs

- Facilitate more inclusive and culturally competent health systems, now and beyond this pandemic
- Develop evidence-based inter-sectoral policies and strategies designed to improve the overall health and social conditions of migrants and respect the rights of migrants to basic human security in host countries
- Facilitate meaningful change to support the inclusion of migrants in host health systems, in alignment with the principles of Universal Health Coverage and the UN Sustainable Development Goals to leave no-one behind

## Data Availability

Datasets are available on request from the authors.

## Contributions

SH conceived the idea for the review and ran the literature searches. SEH, SH, and CC did the abstract and full text screening, data extraction, and synthesis. AD and CC did the quality appraisal. AFC, MO, MN, AR, CG, KB, AV, FW, IC-M, FS, and BN provided national datasets and grey literature. SEH and SH wrote a first draft of the paper with input from all authors.

## Acknowledgements

SEH and AD are supported by Medical Research Council PhD studentships (MR/N013638/1). SH is funded by the NIHR (NIHR Advanced Fellowship NIHR300072) for this research project. The views expressed in this publication are those of the author(s) and not necessarily those of the NIHR, NHS or the UK Department of Health and Social Care. SH is also funded by the Academy of Medical Sciences (SBF005\1111), and by the European Society of Clinical Microbiology and Infectious Diseases (ESCMID) through a joint ESCMID Study Group for Infections in Travellers and Migrants (ESGITM) and ESCMID Study Group for Mycobacterial Infections (ESGMYC) Research grant. AFC is funded by the Academy of Medical Sciences (SBF005\1111) and the NIHR (NIHR300072). JC is funded by the NIHR (NIHR in-practice clinical fellowship NIHR300290). MO is supported by an Economic and Social Research Council (ESRC) PhD studentship.

---

**Supplementary Table 1:**
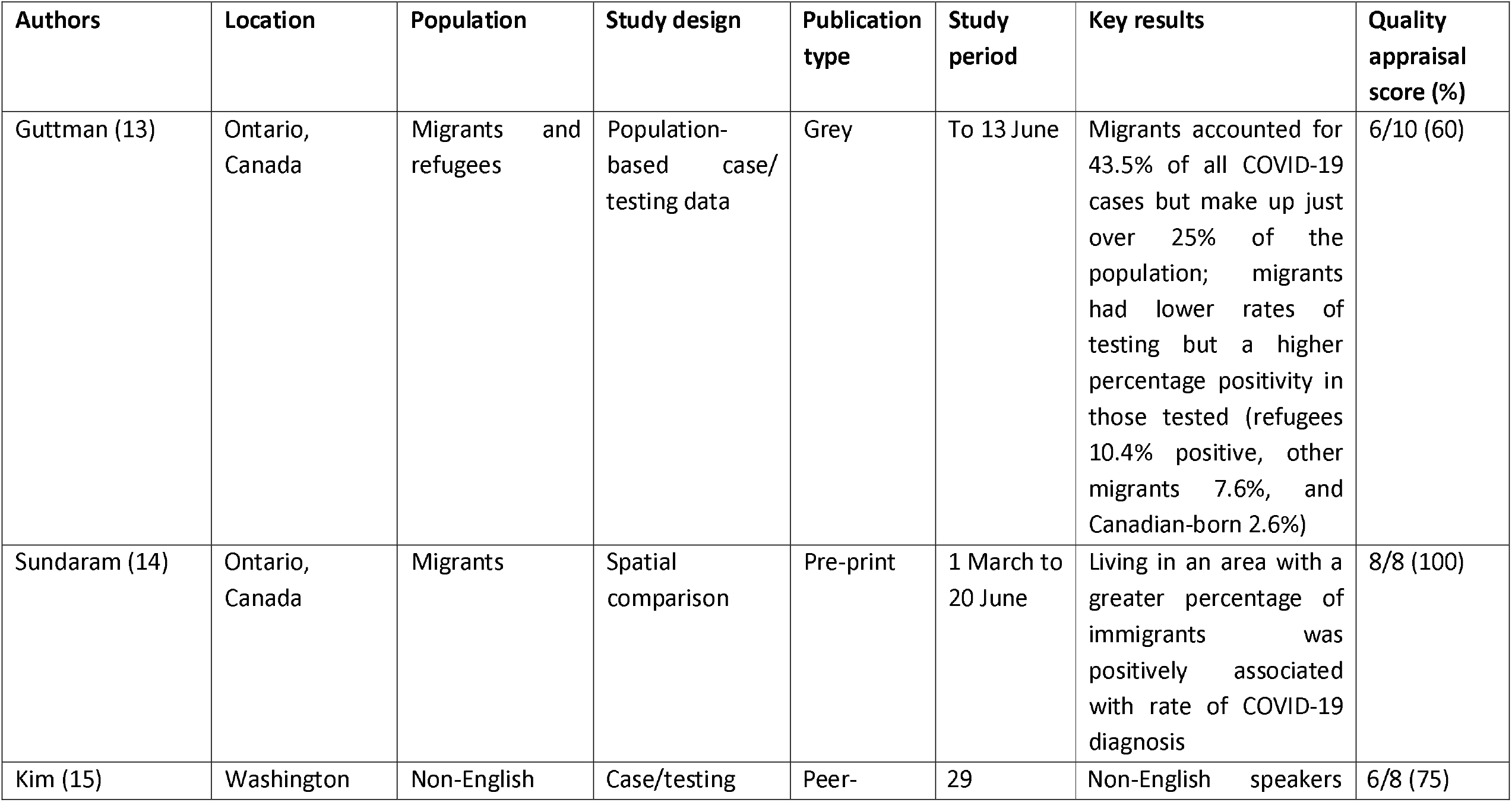
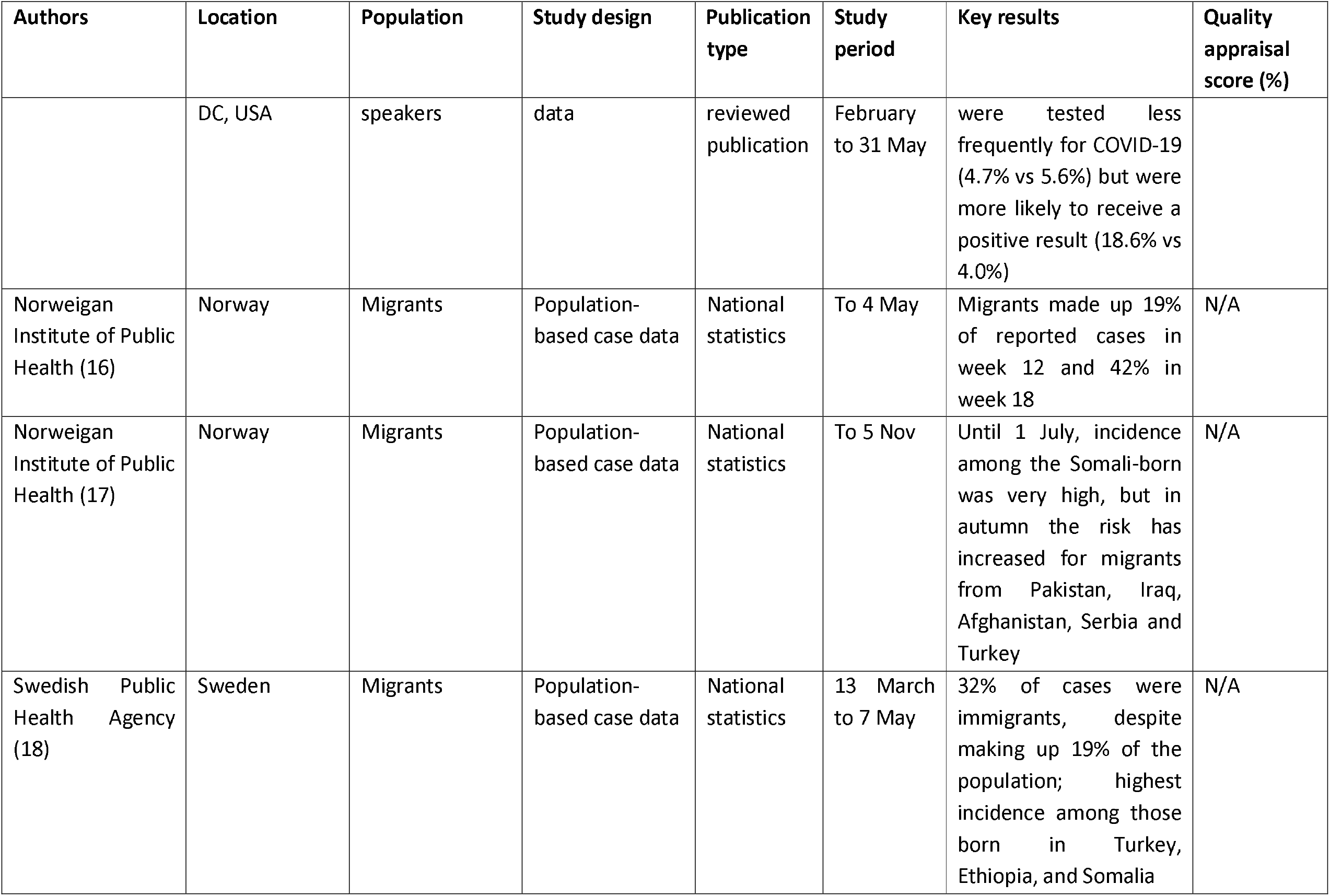
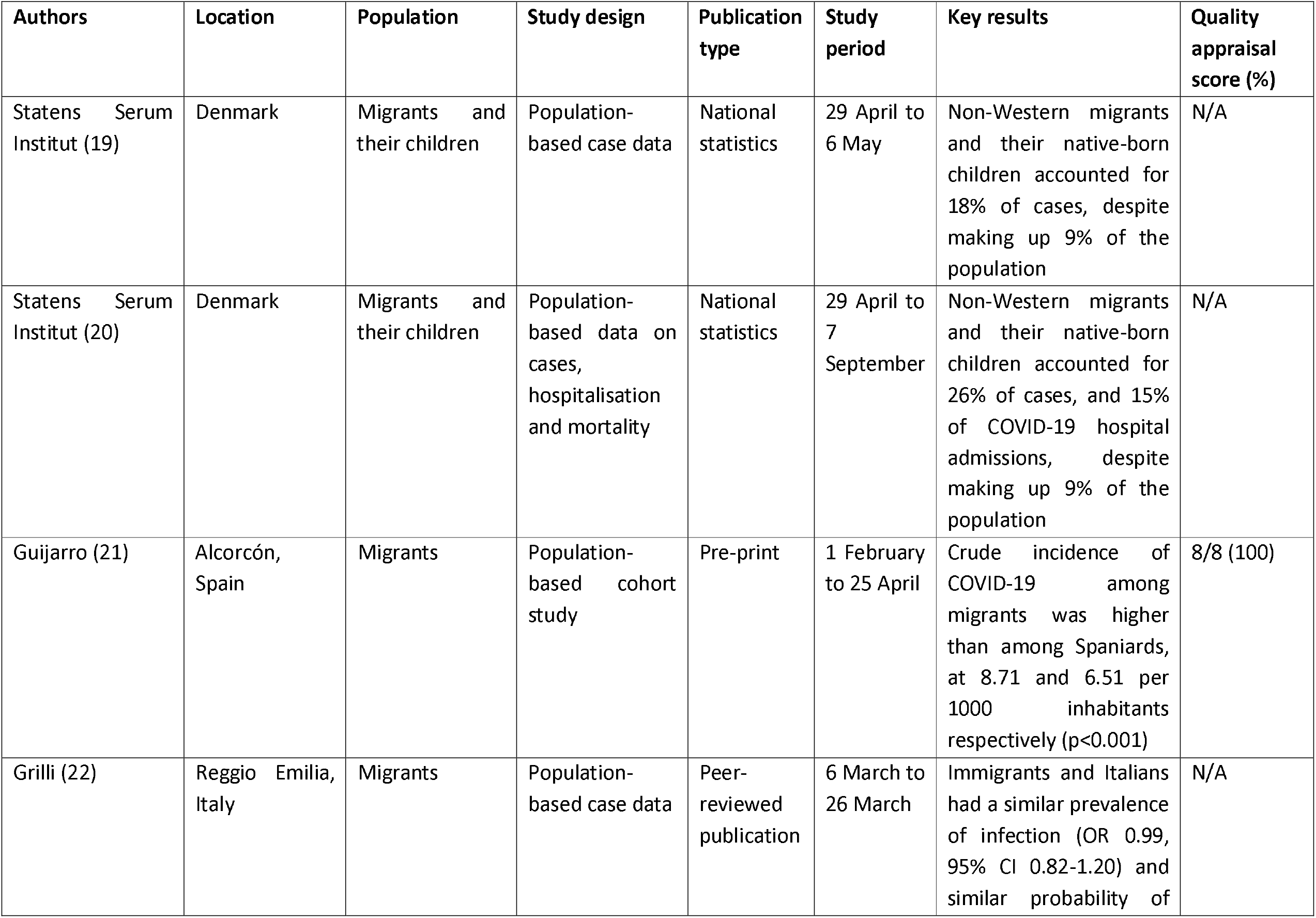
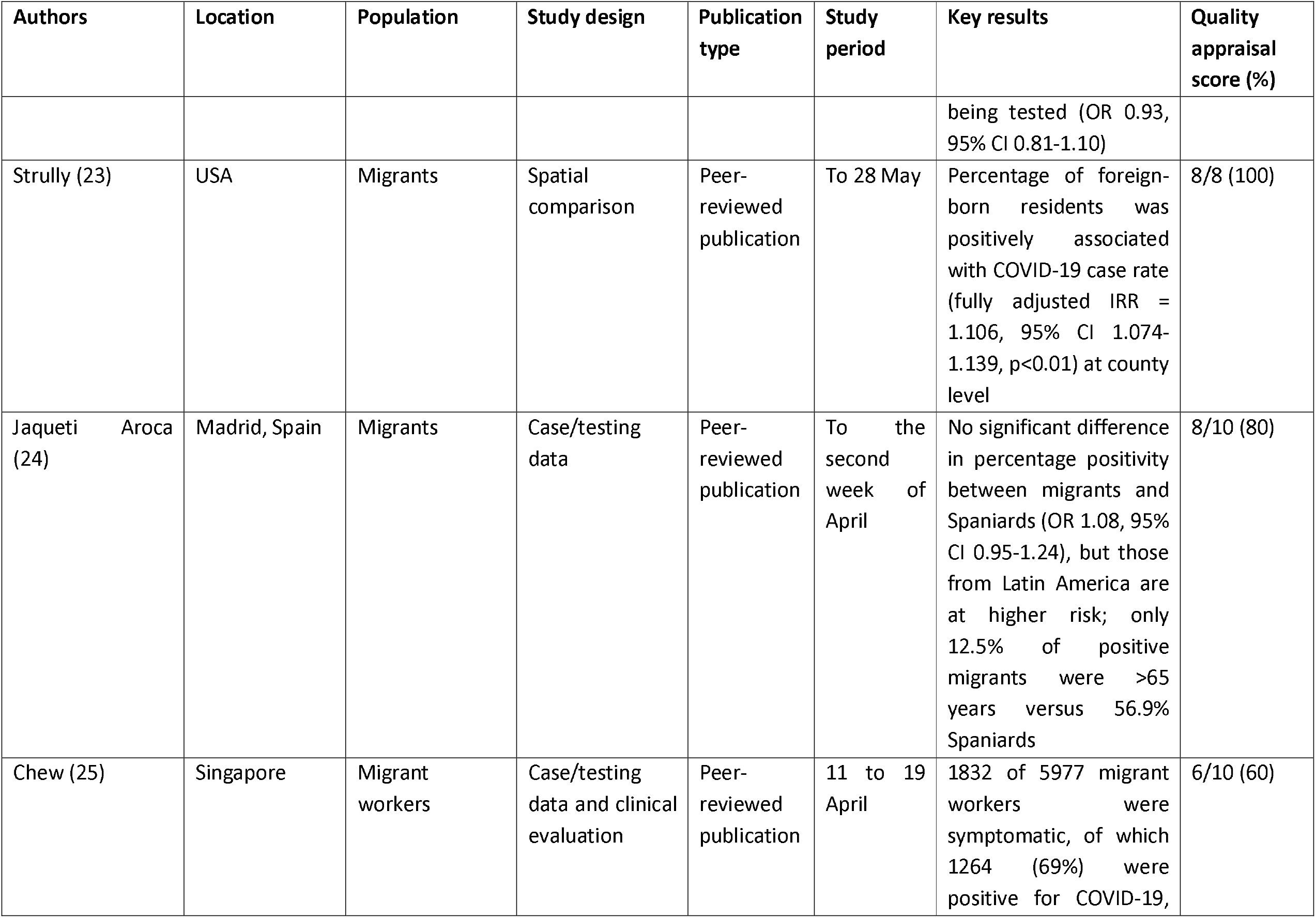
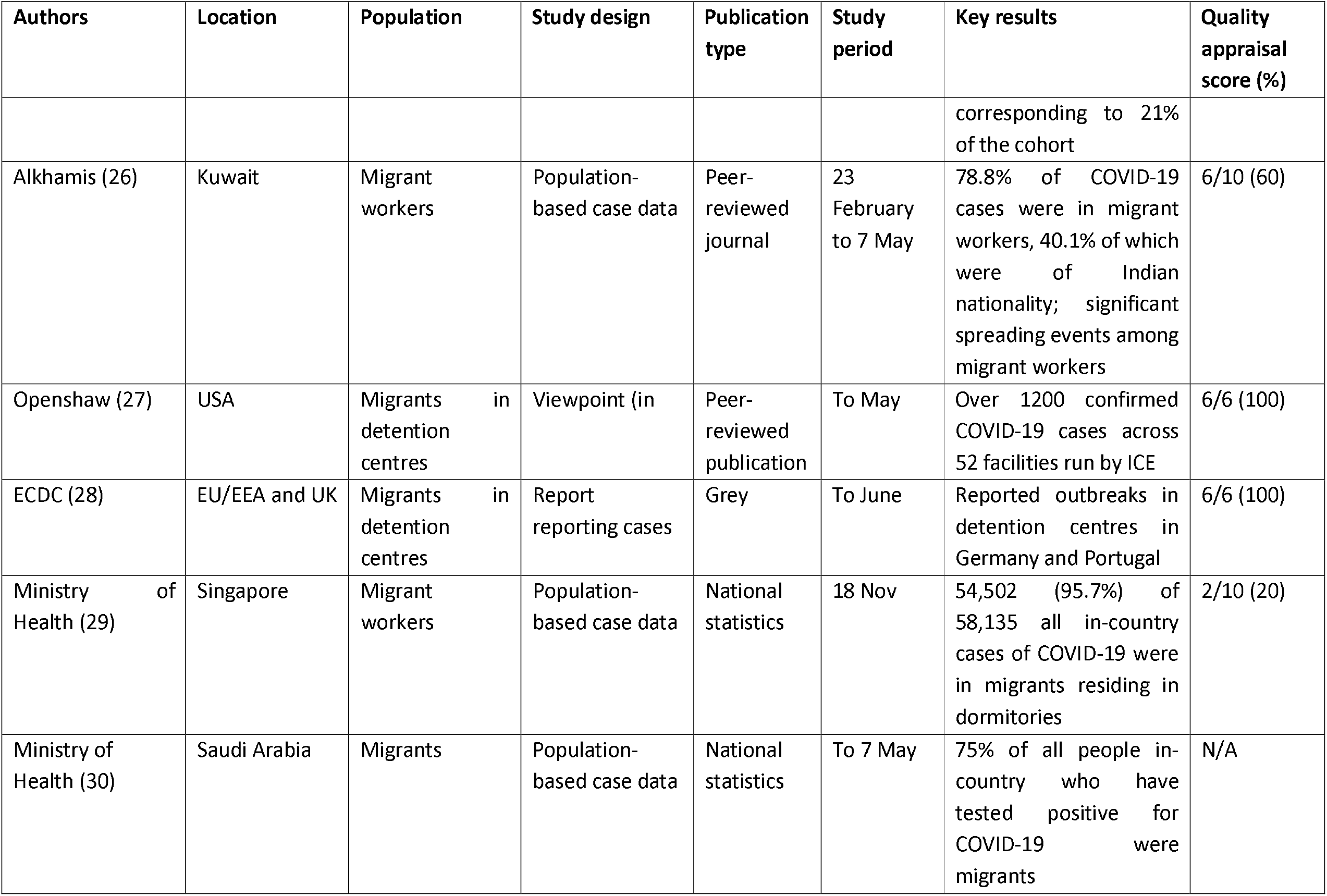
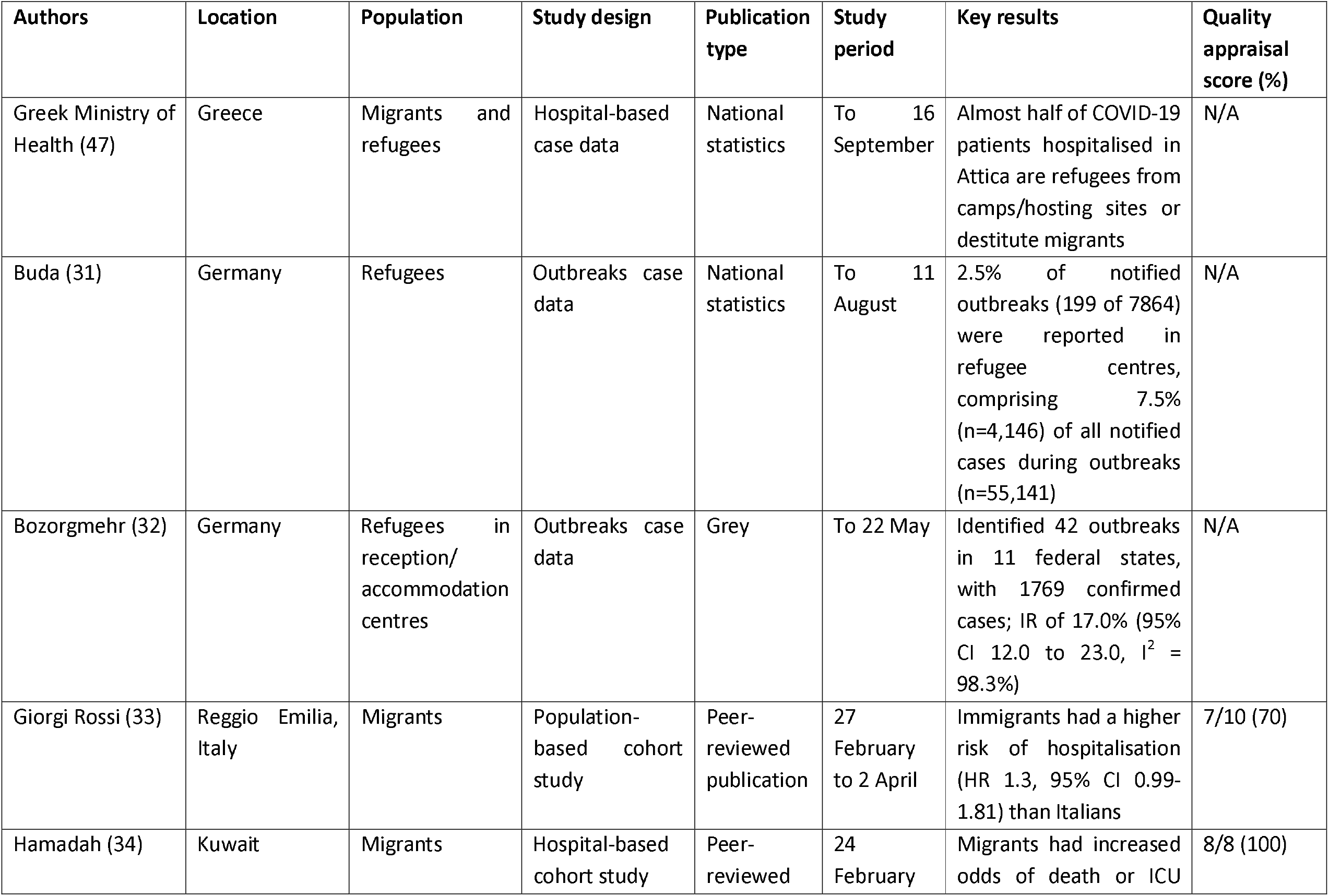
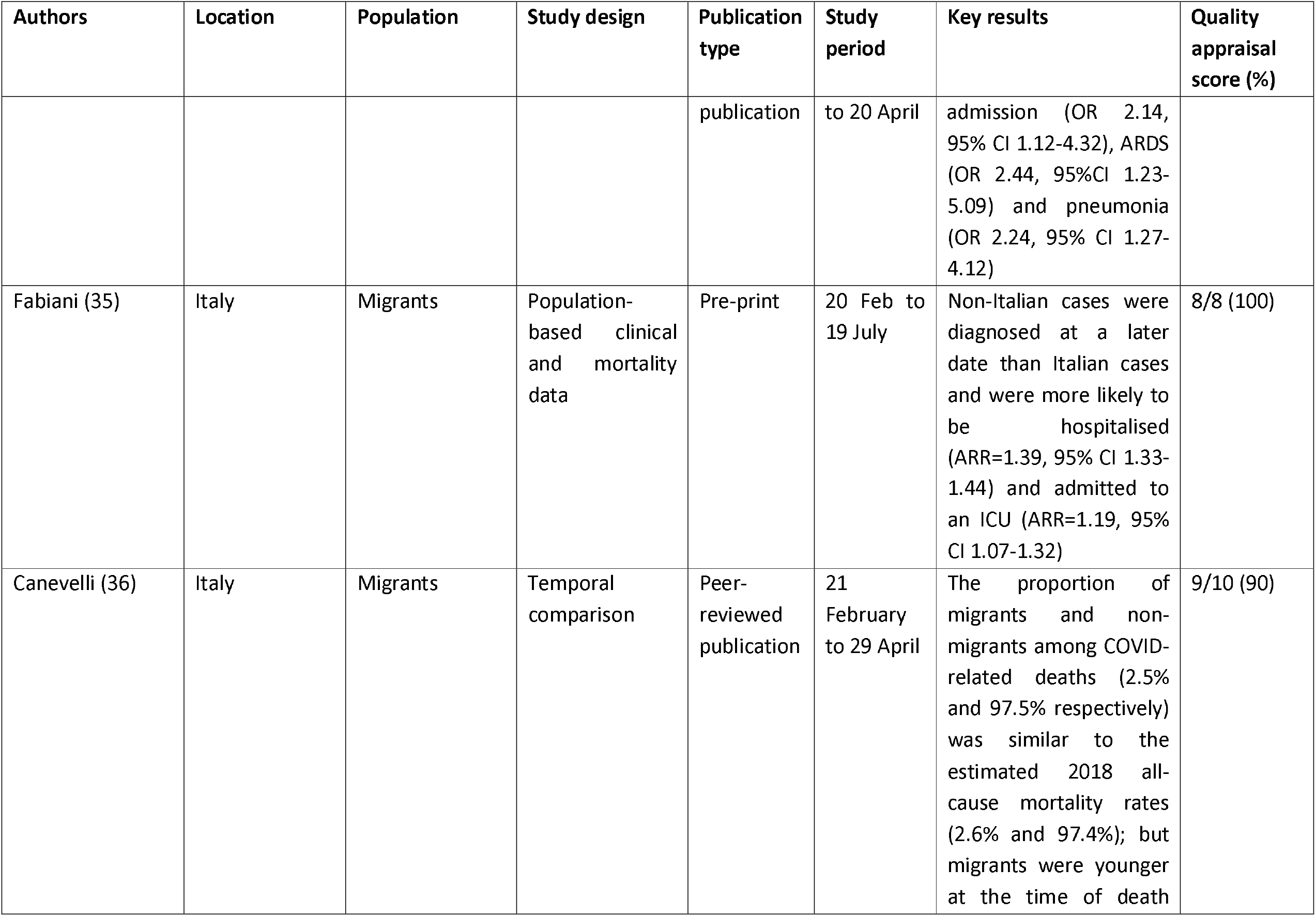
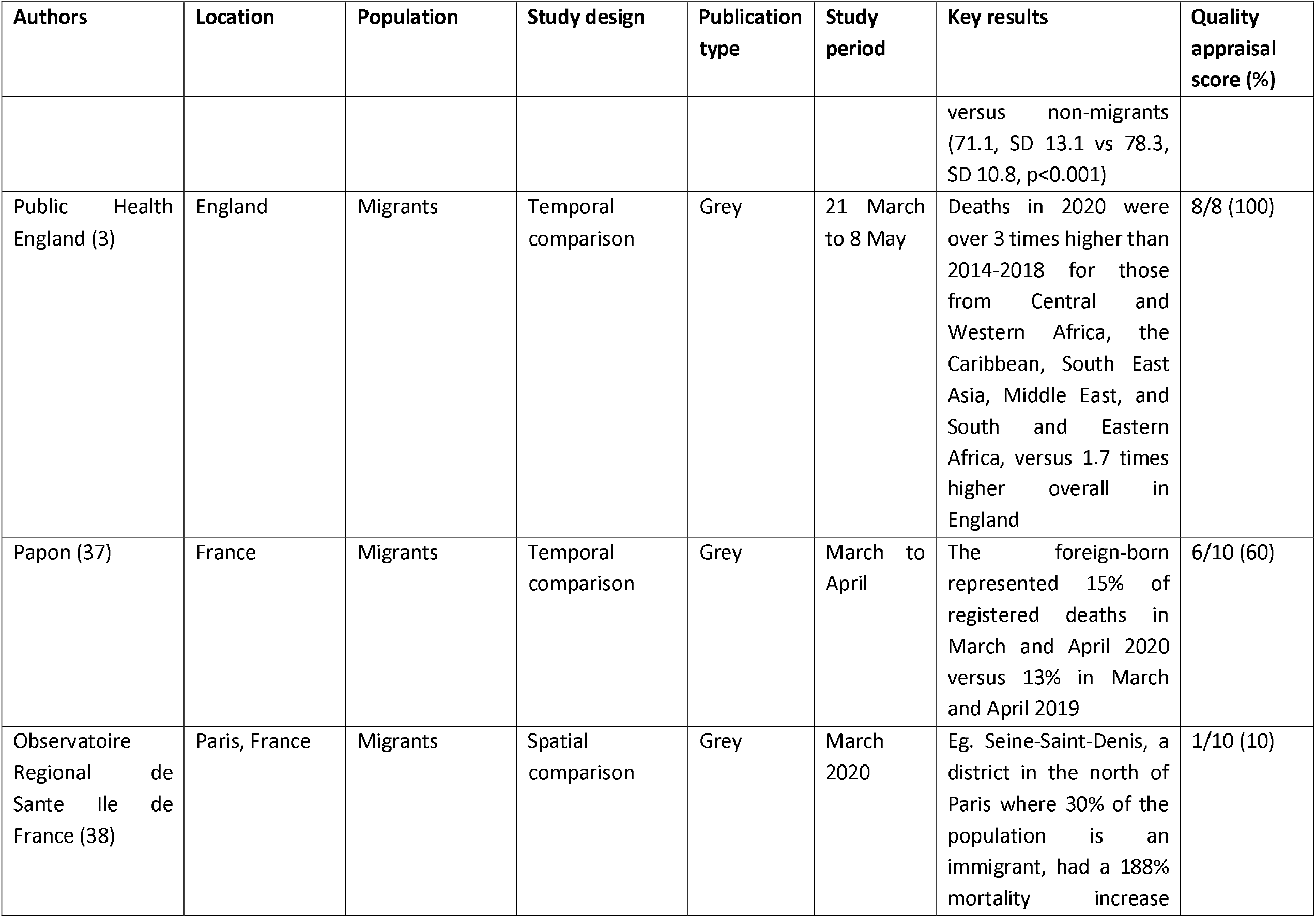
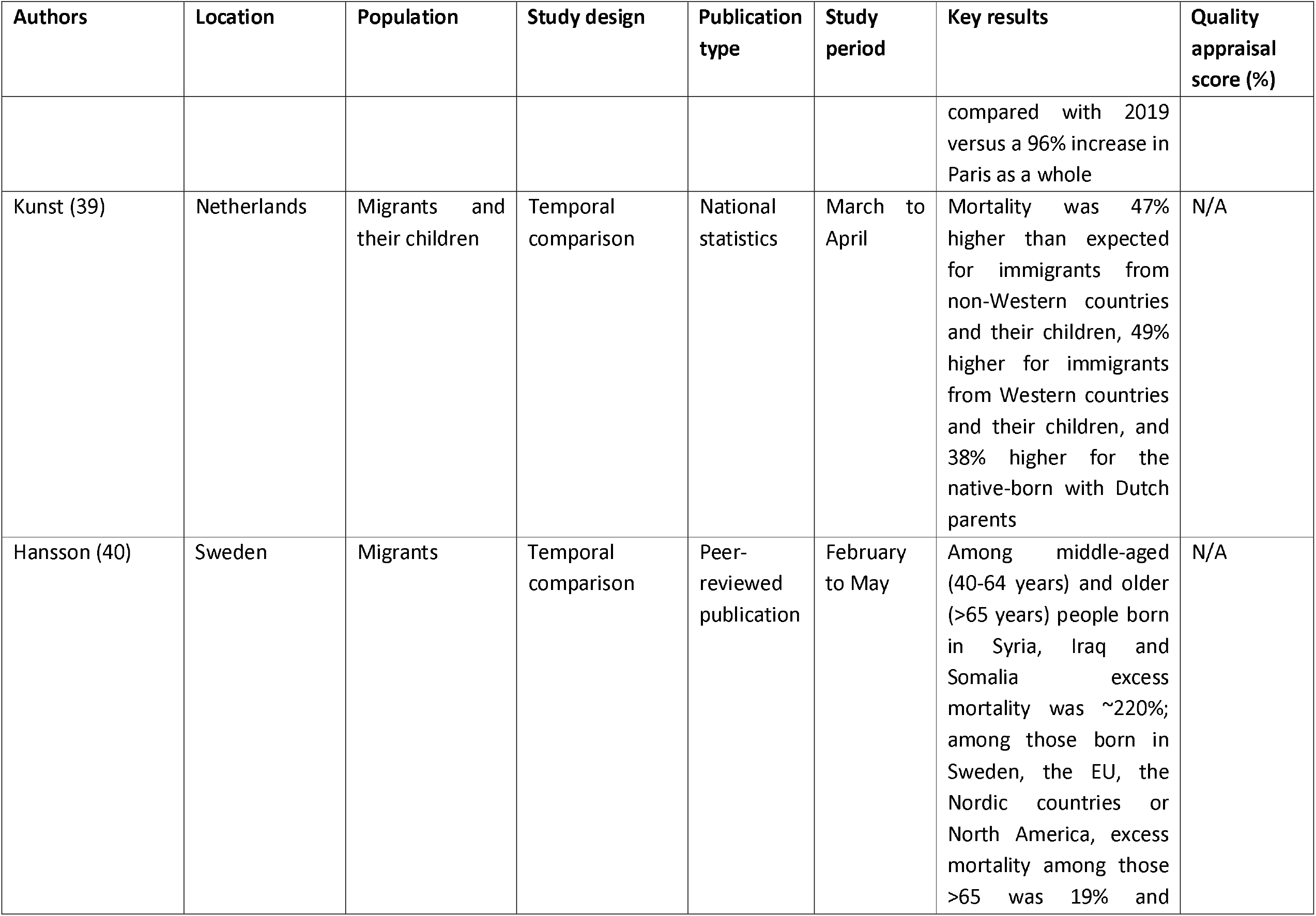
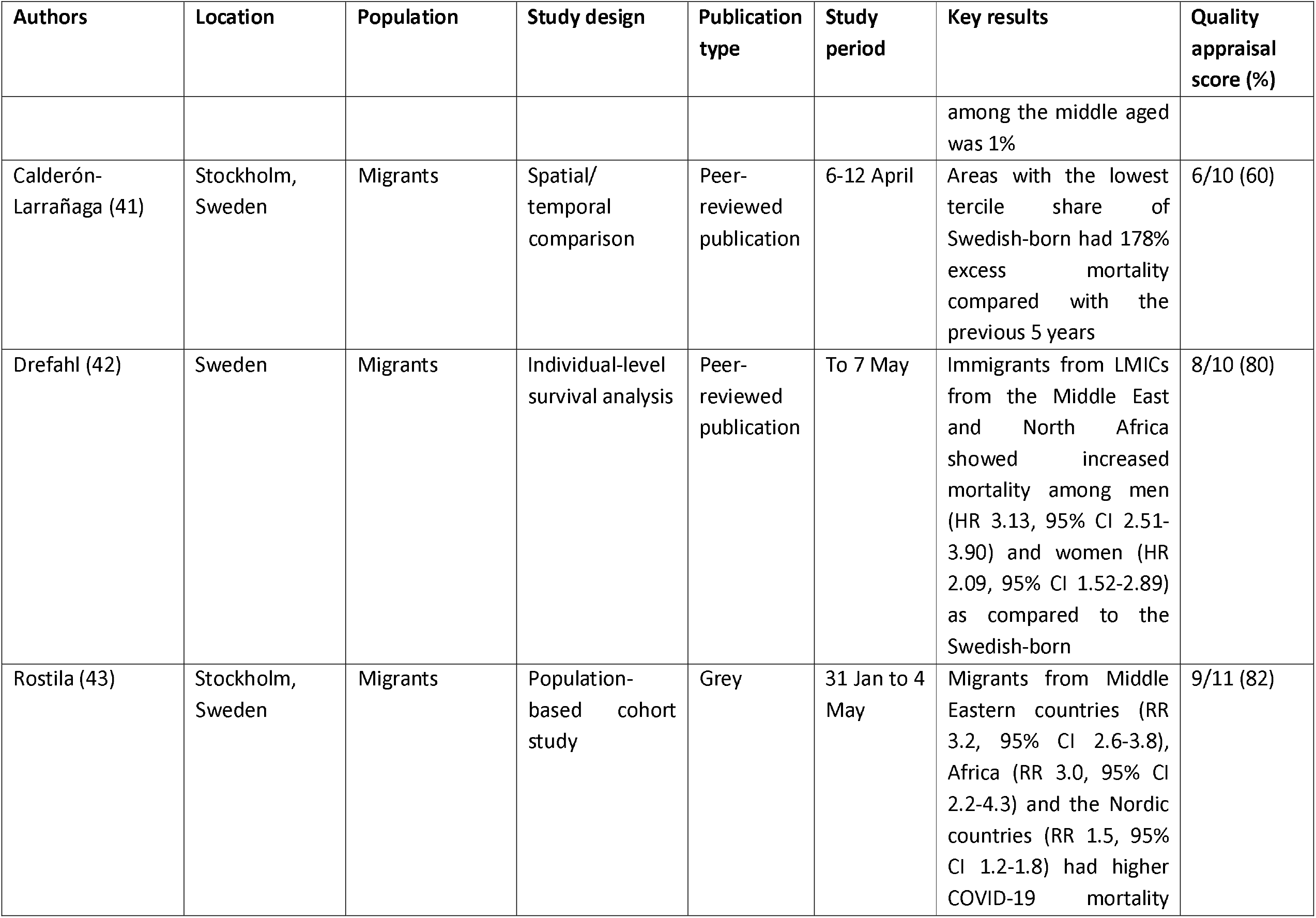
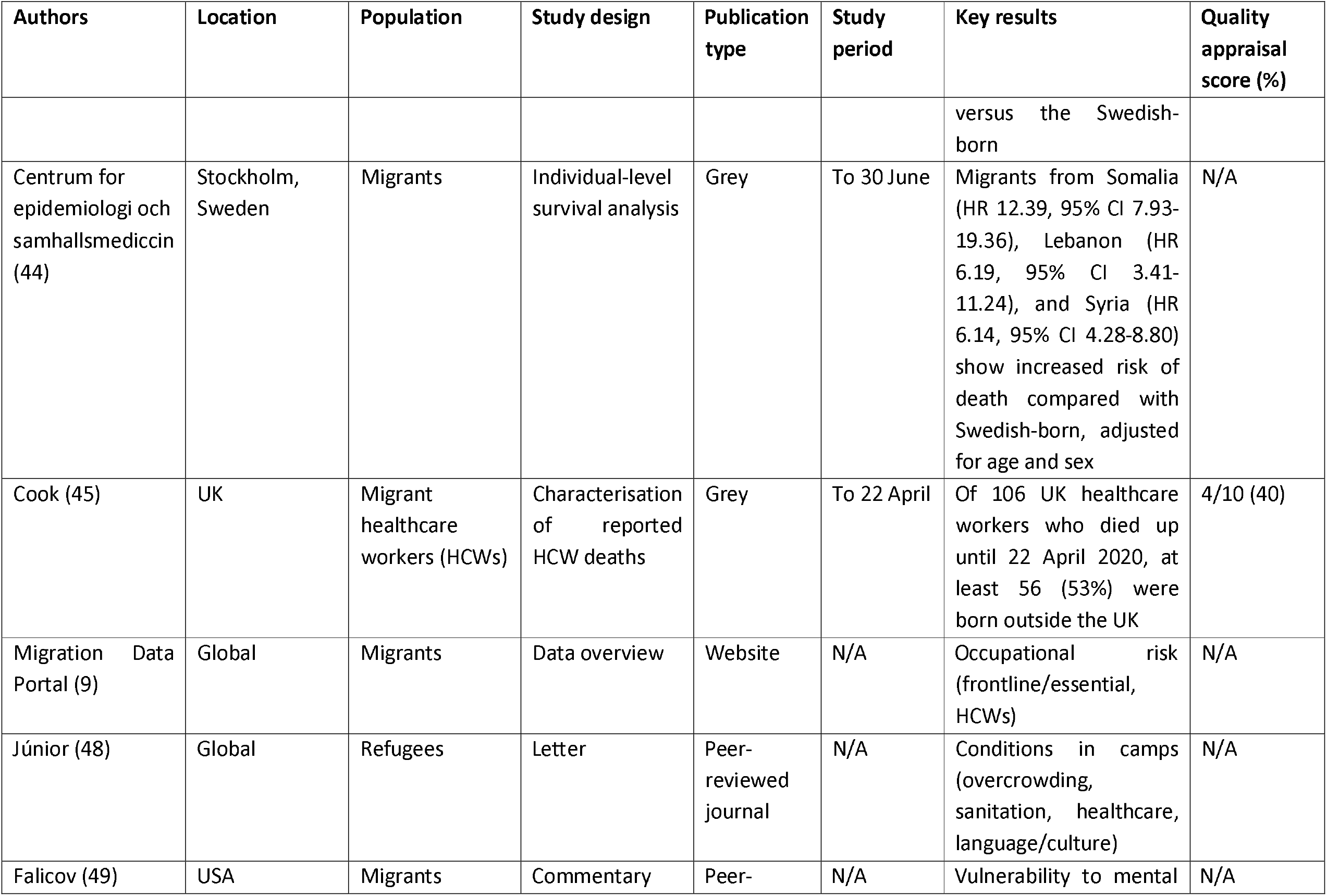
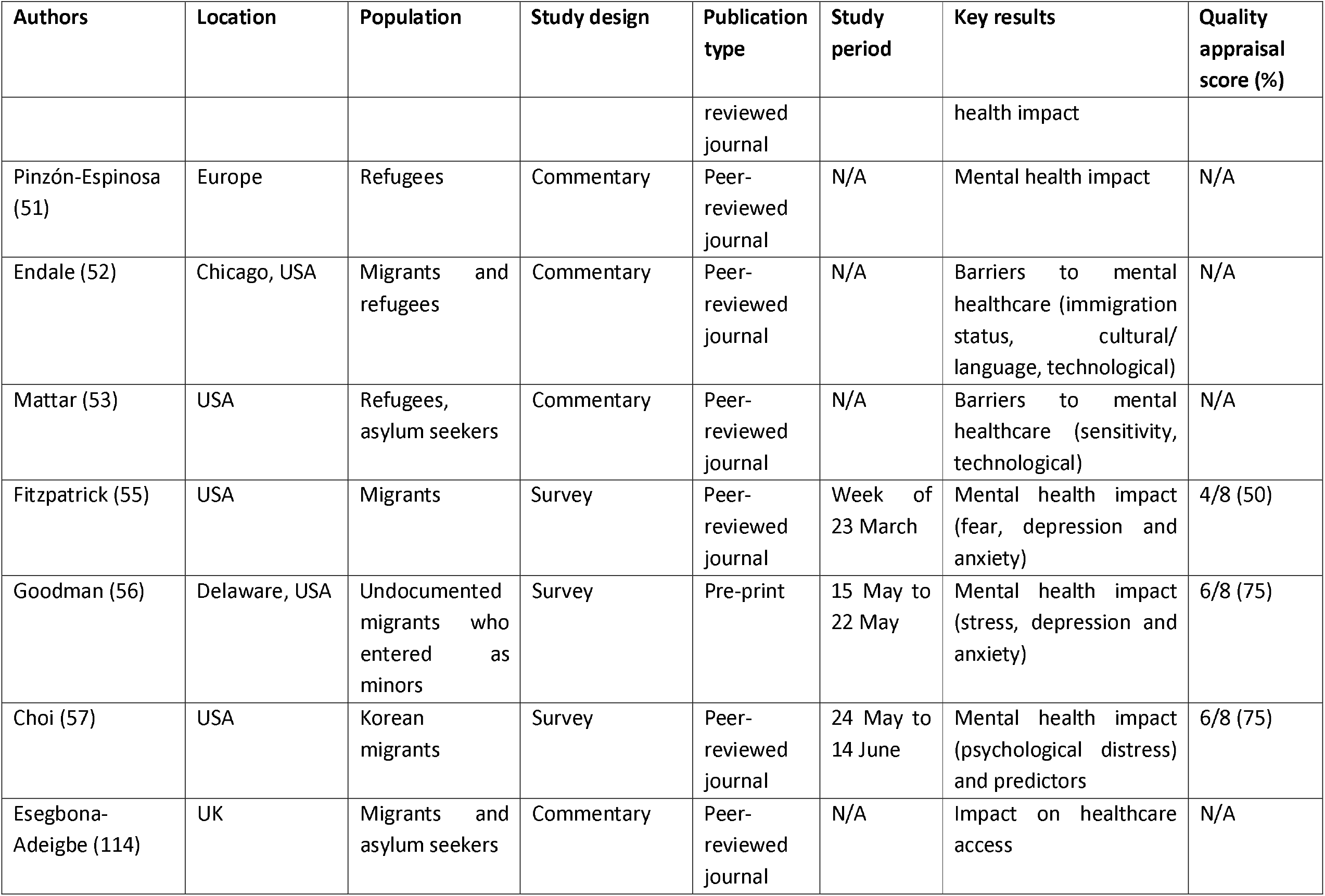
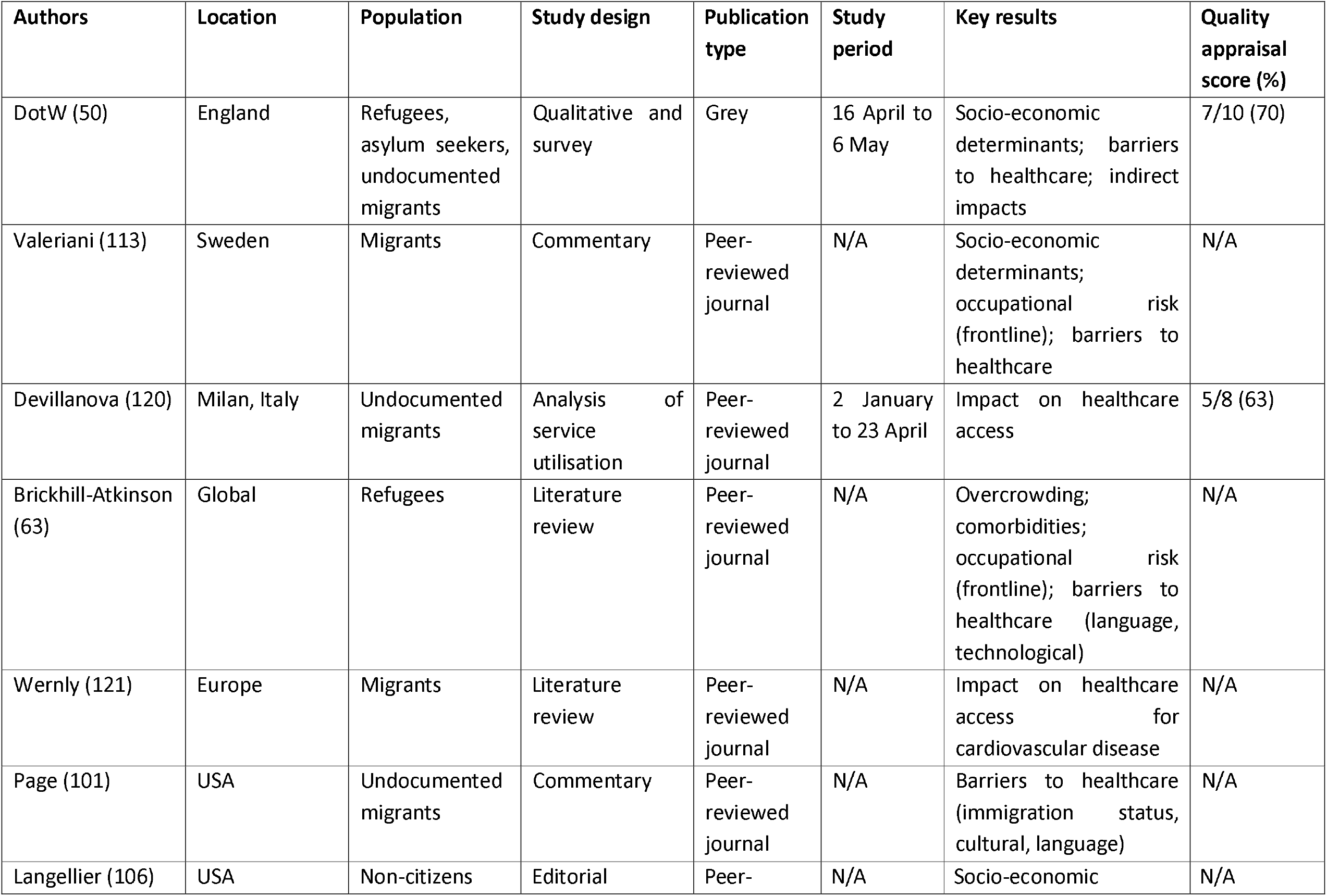
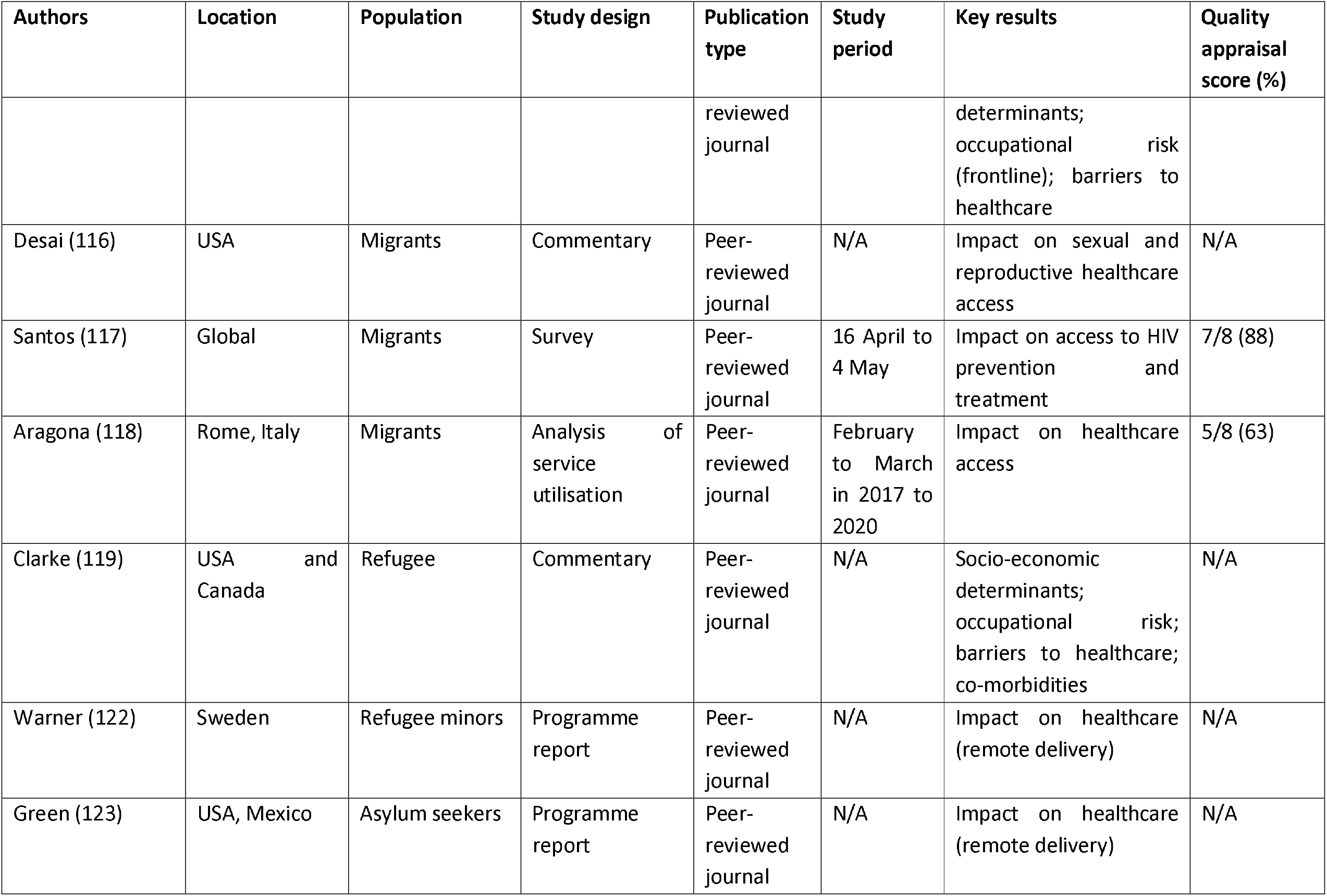
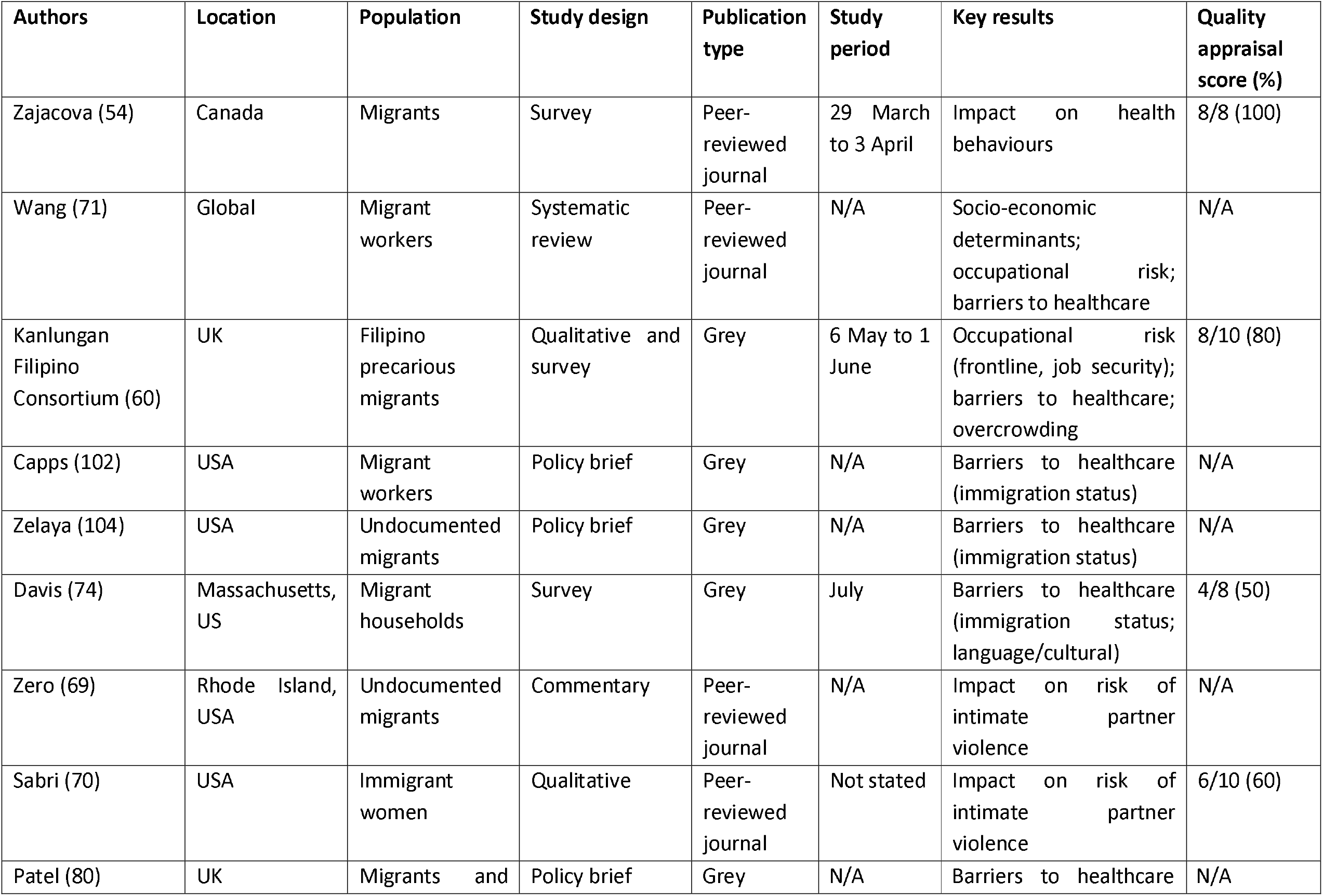
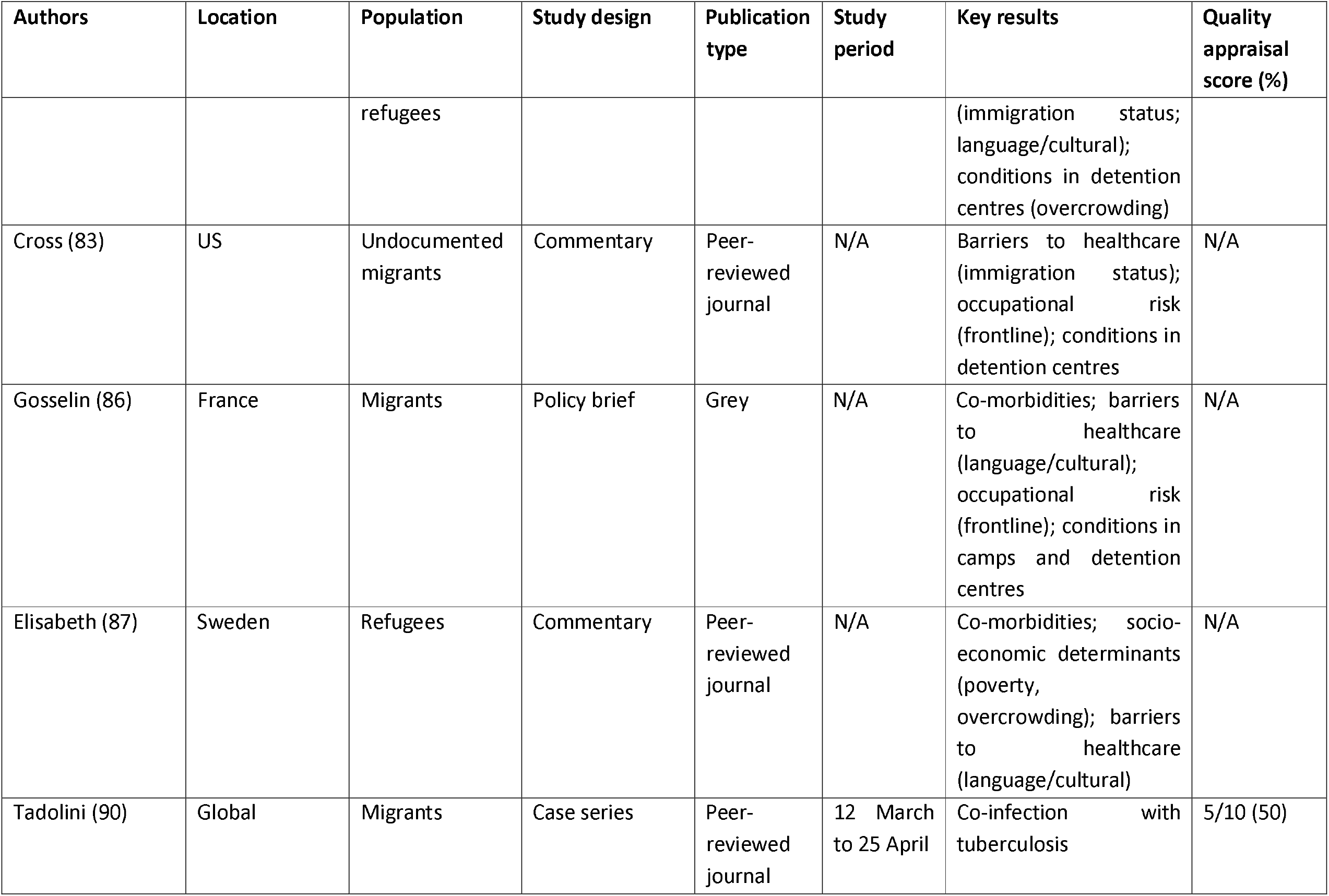
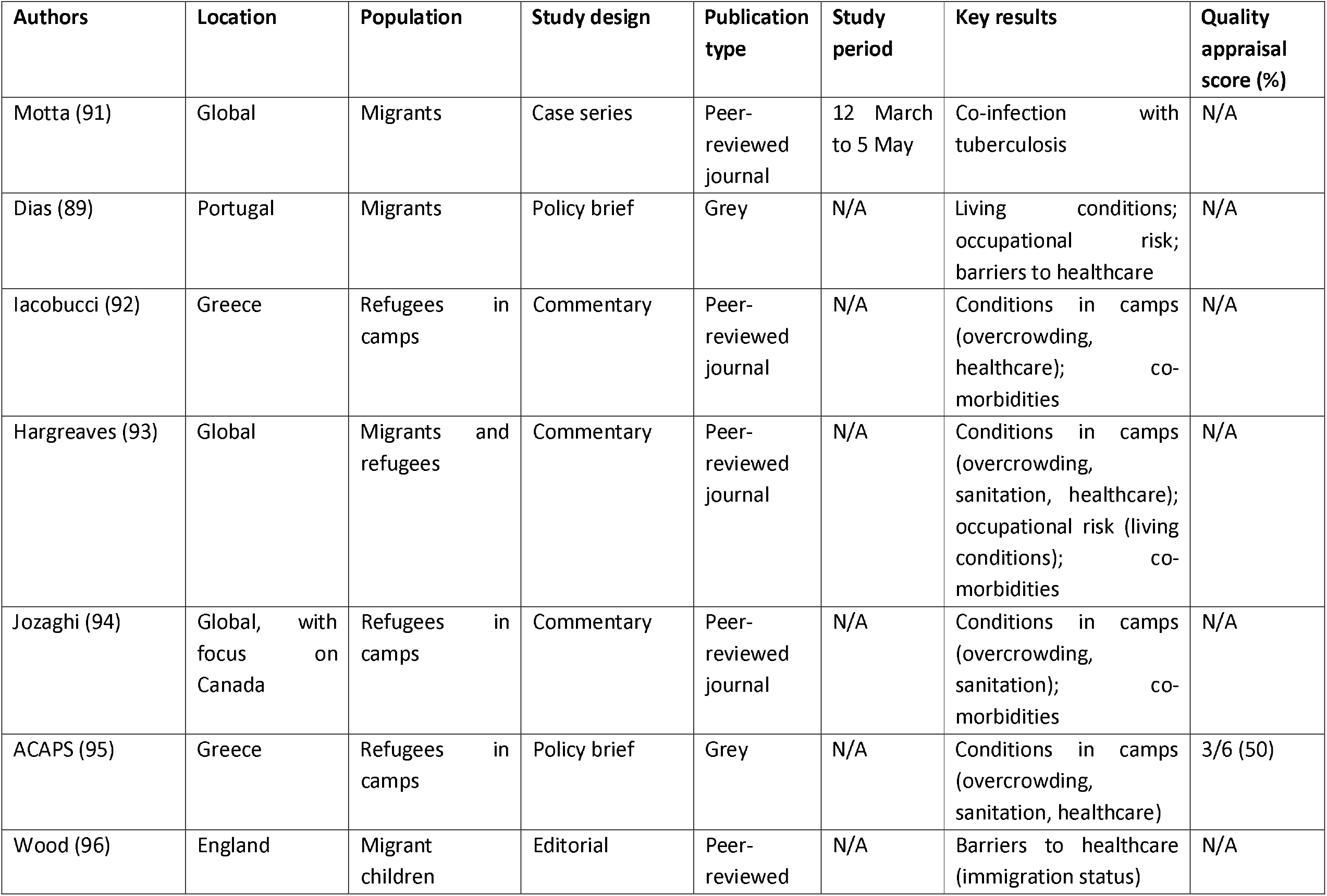
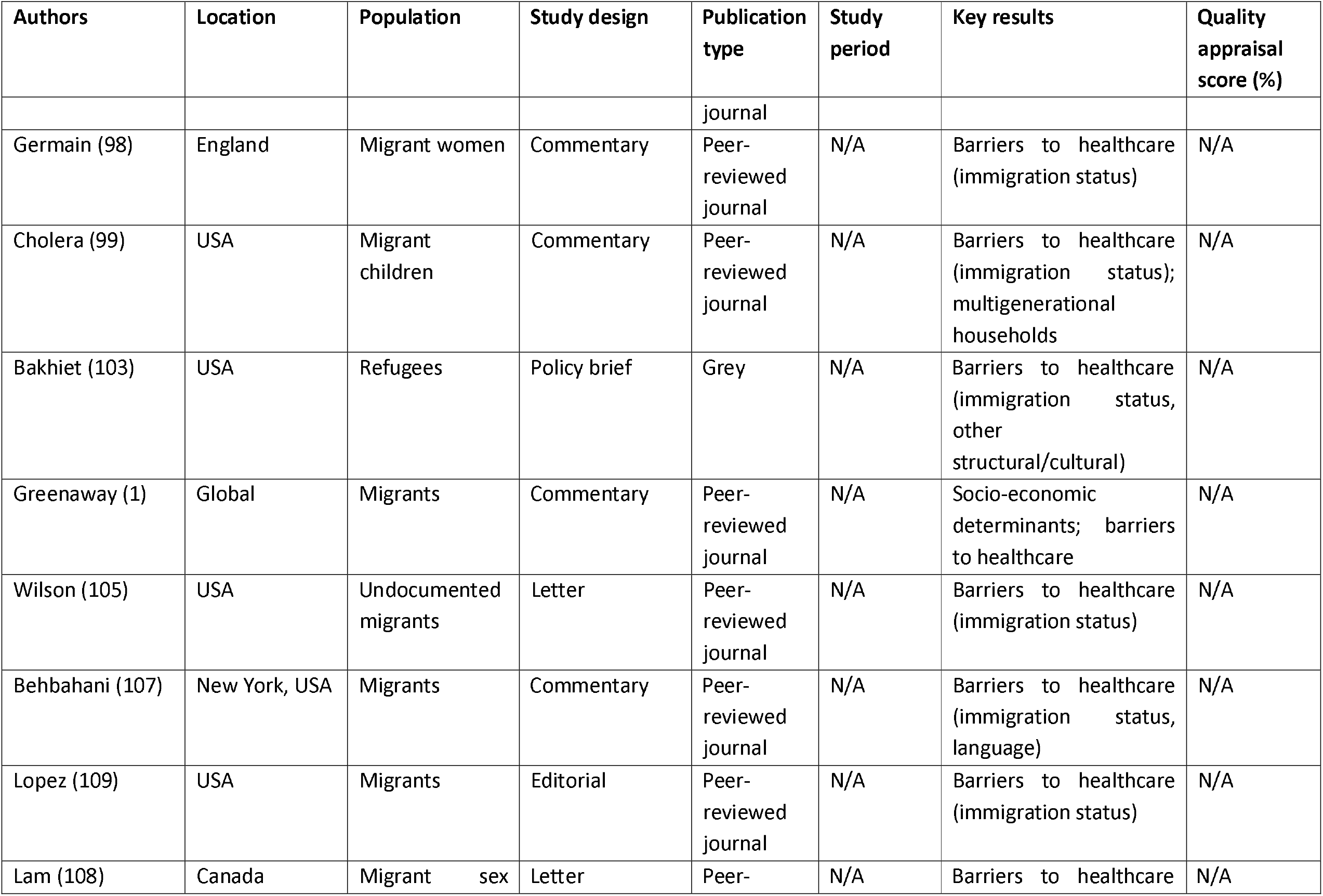
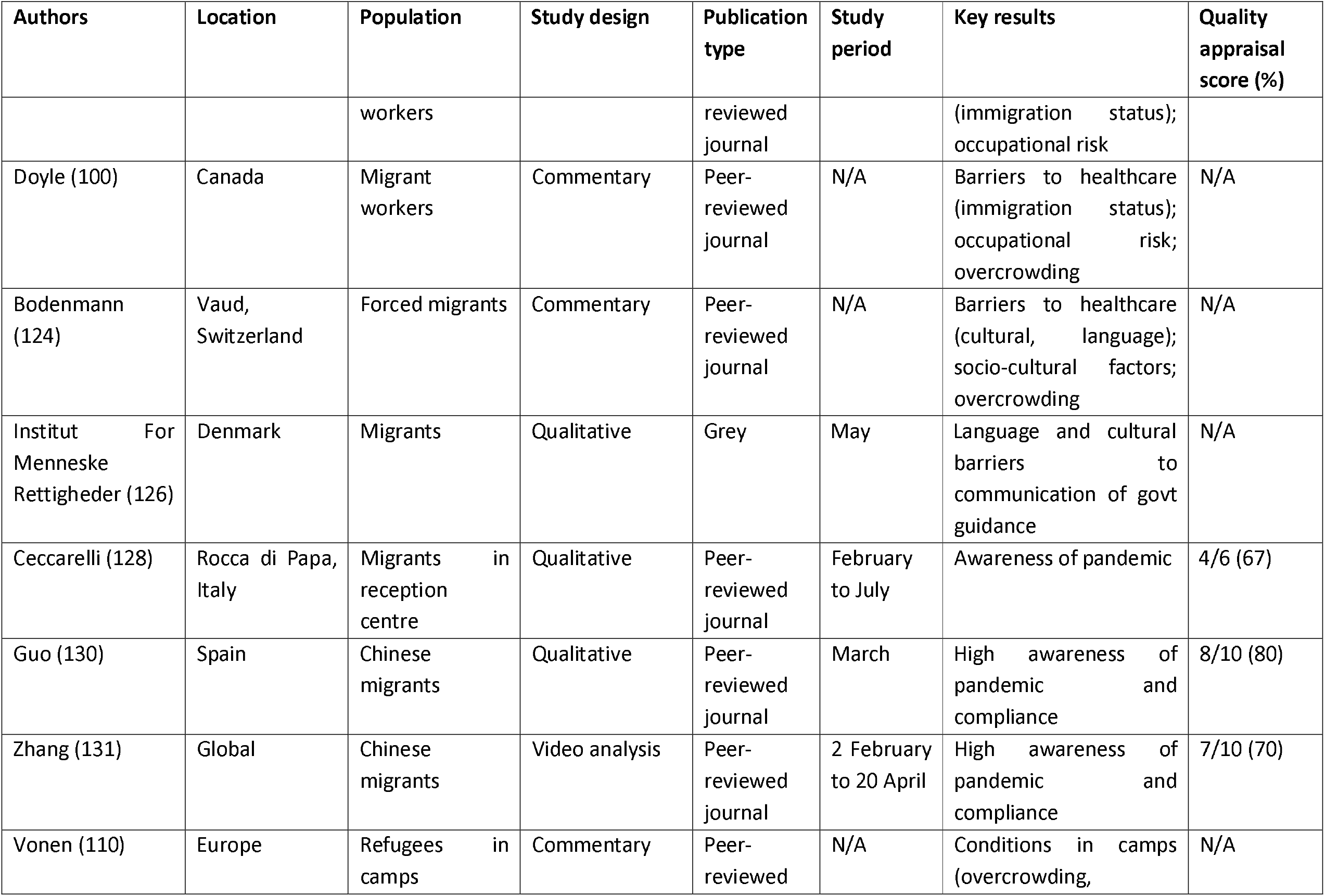
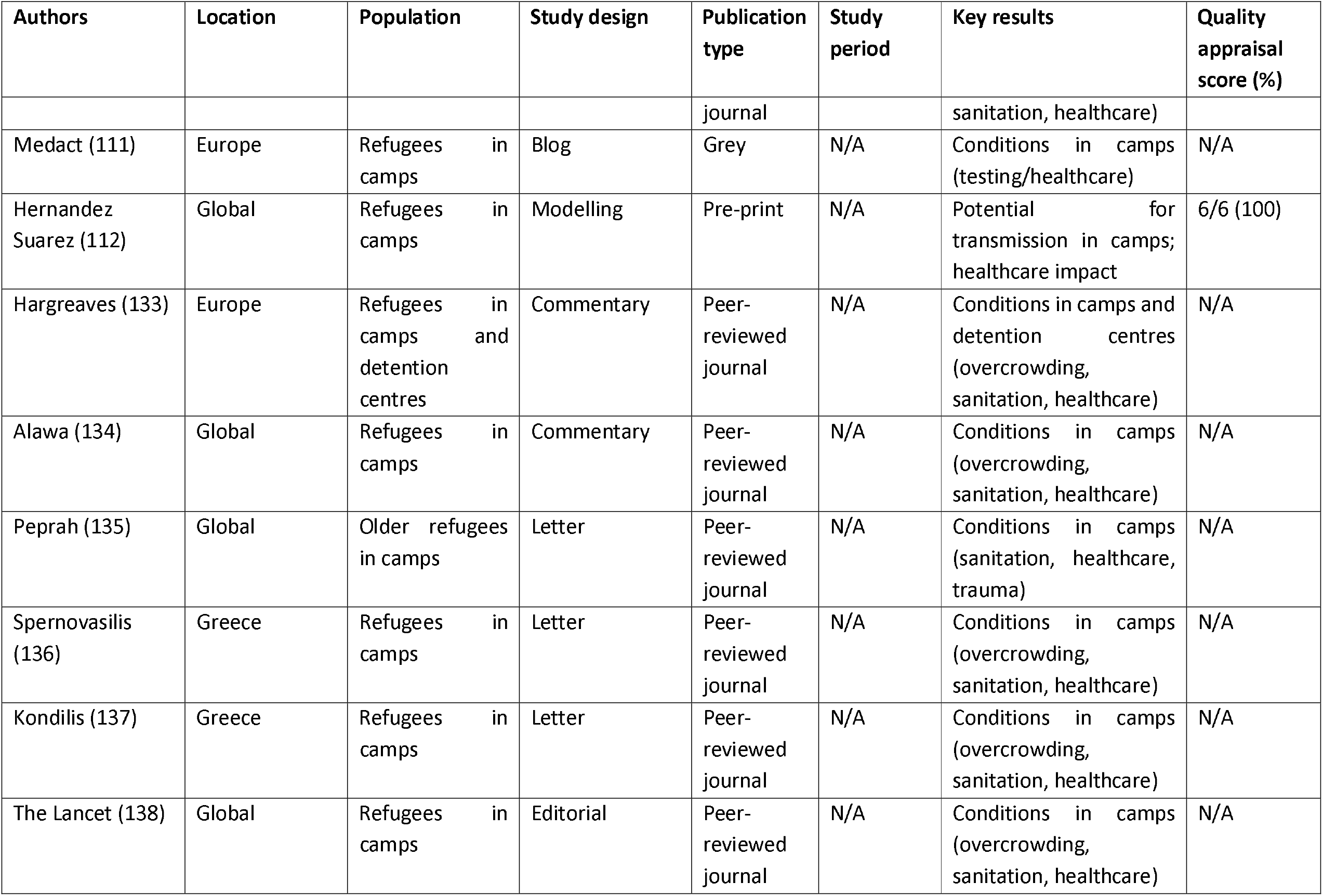
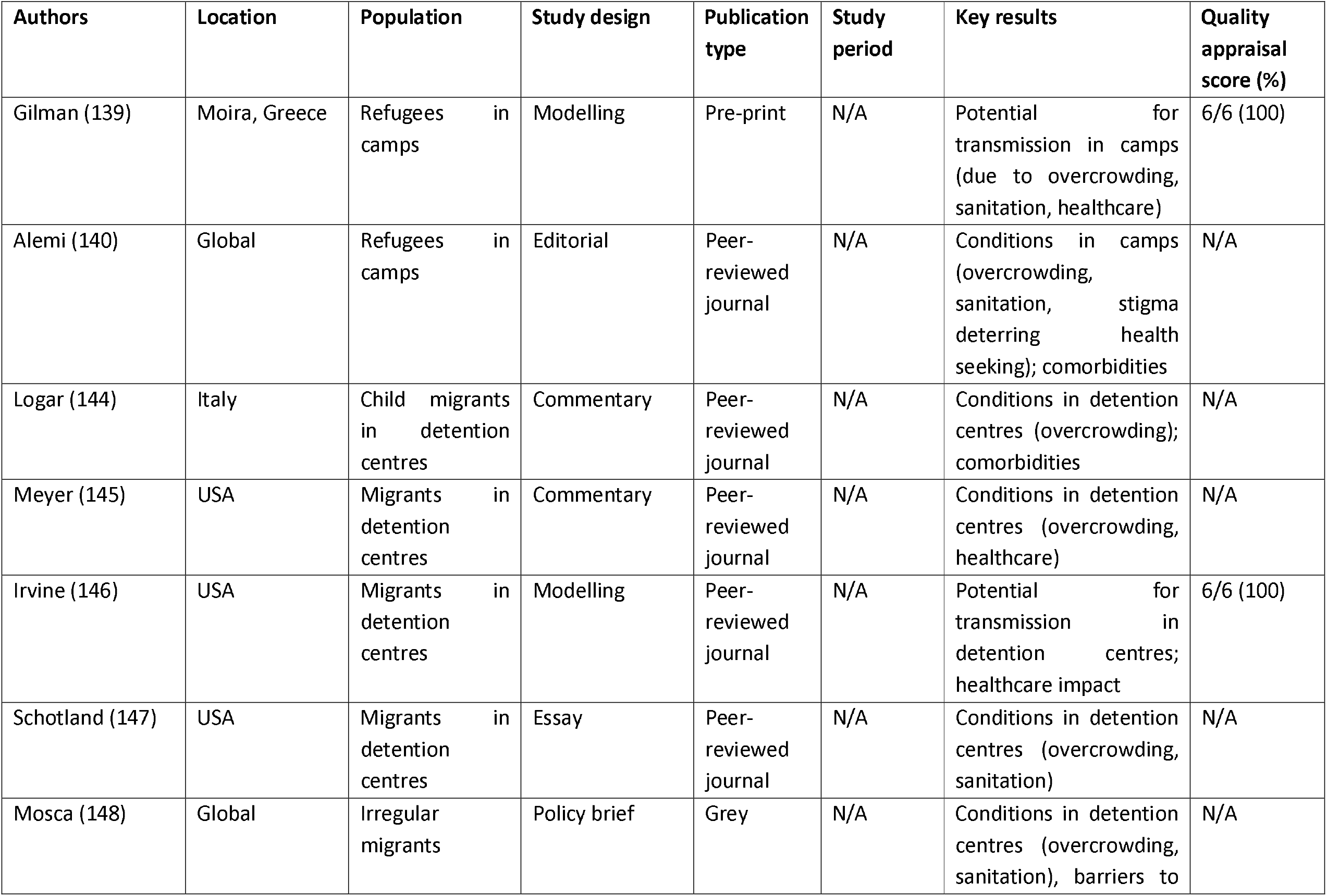
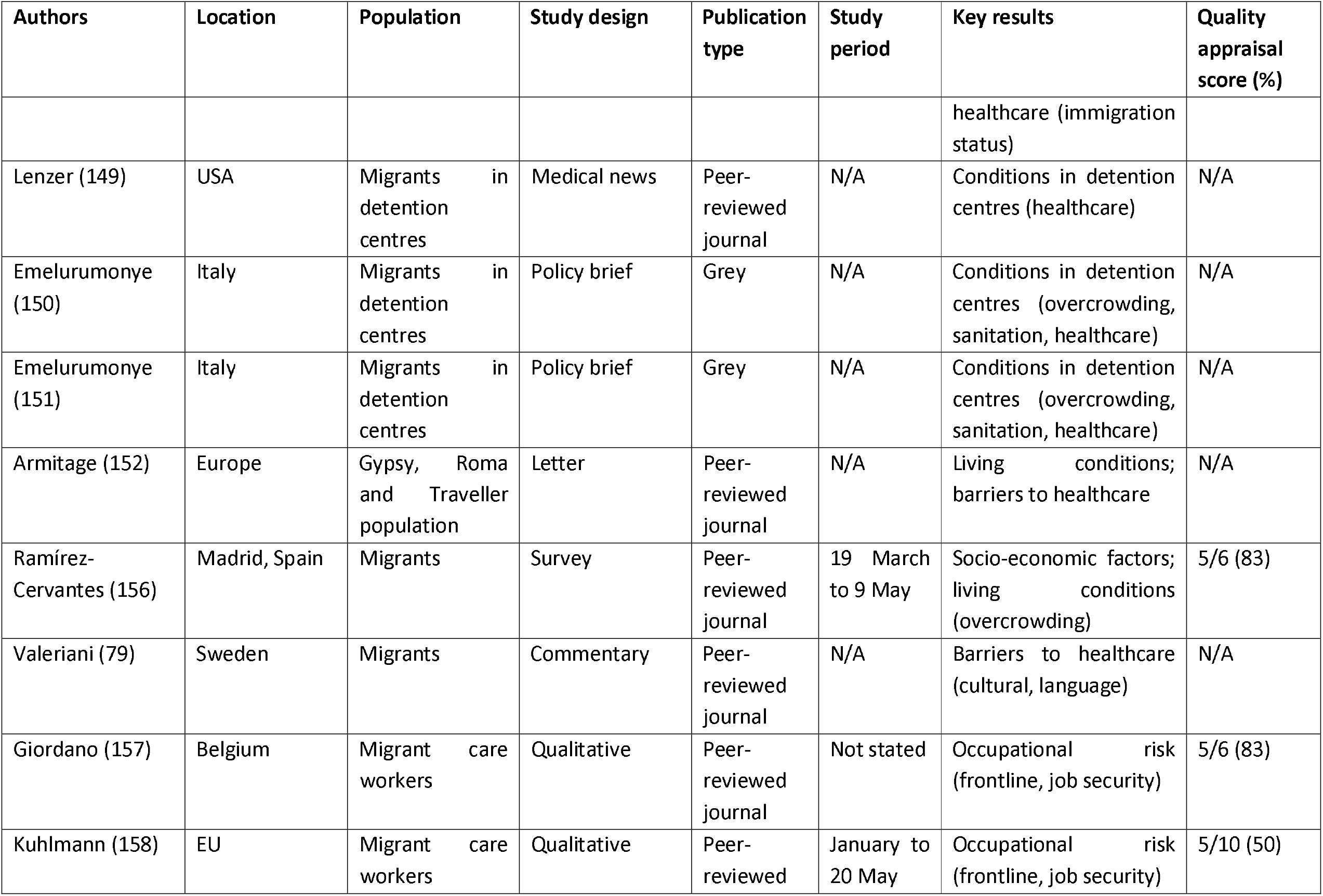
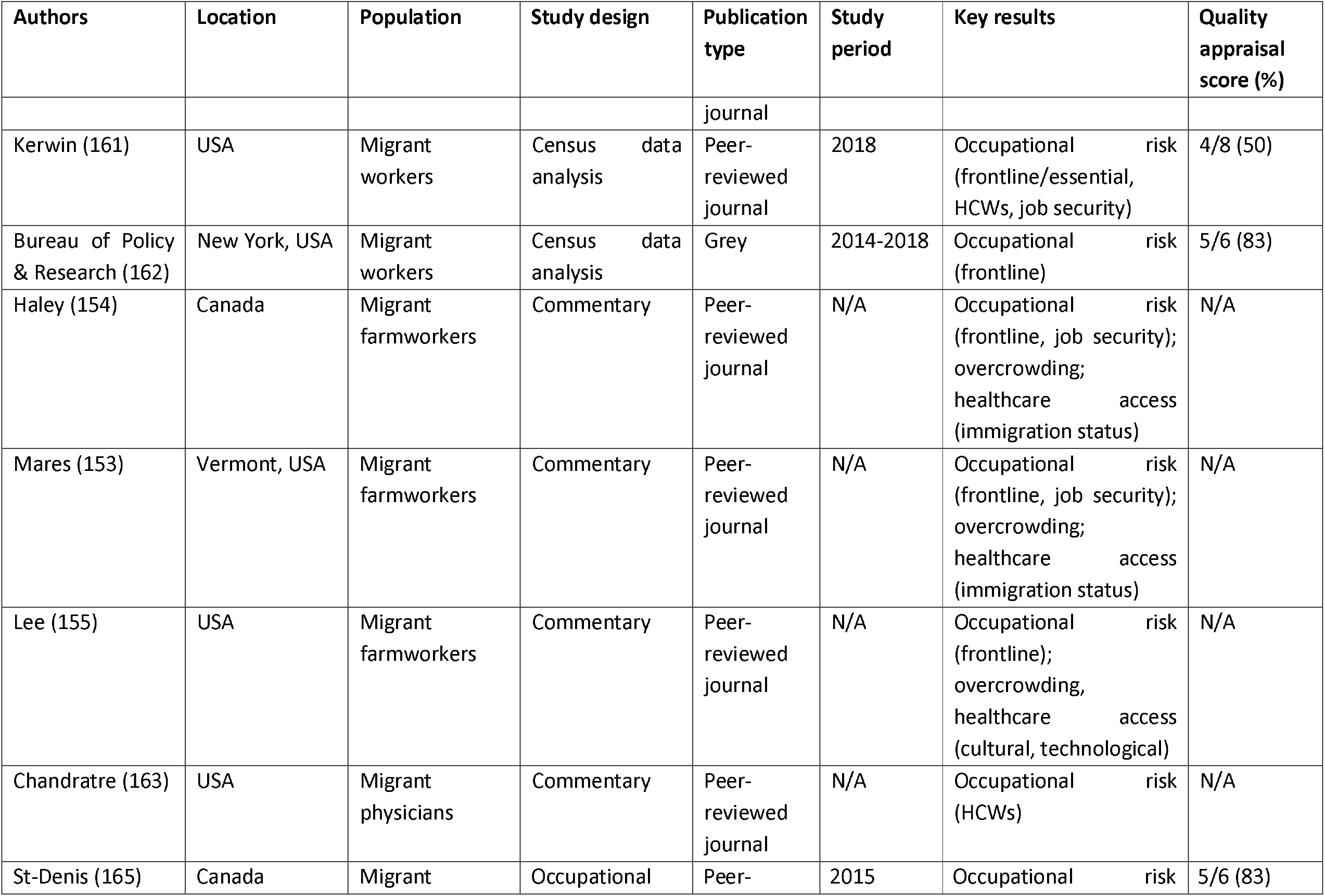
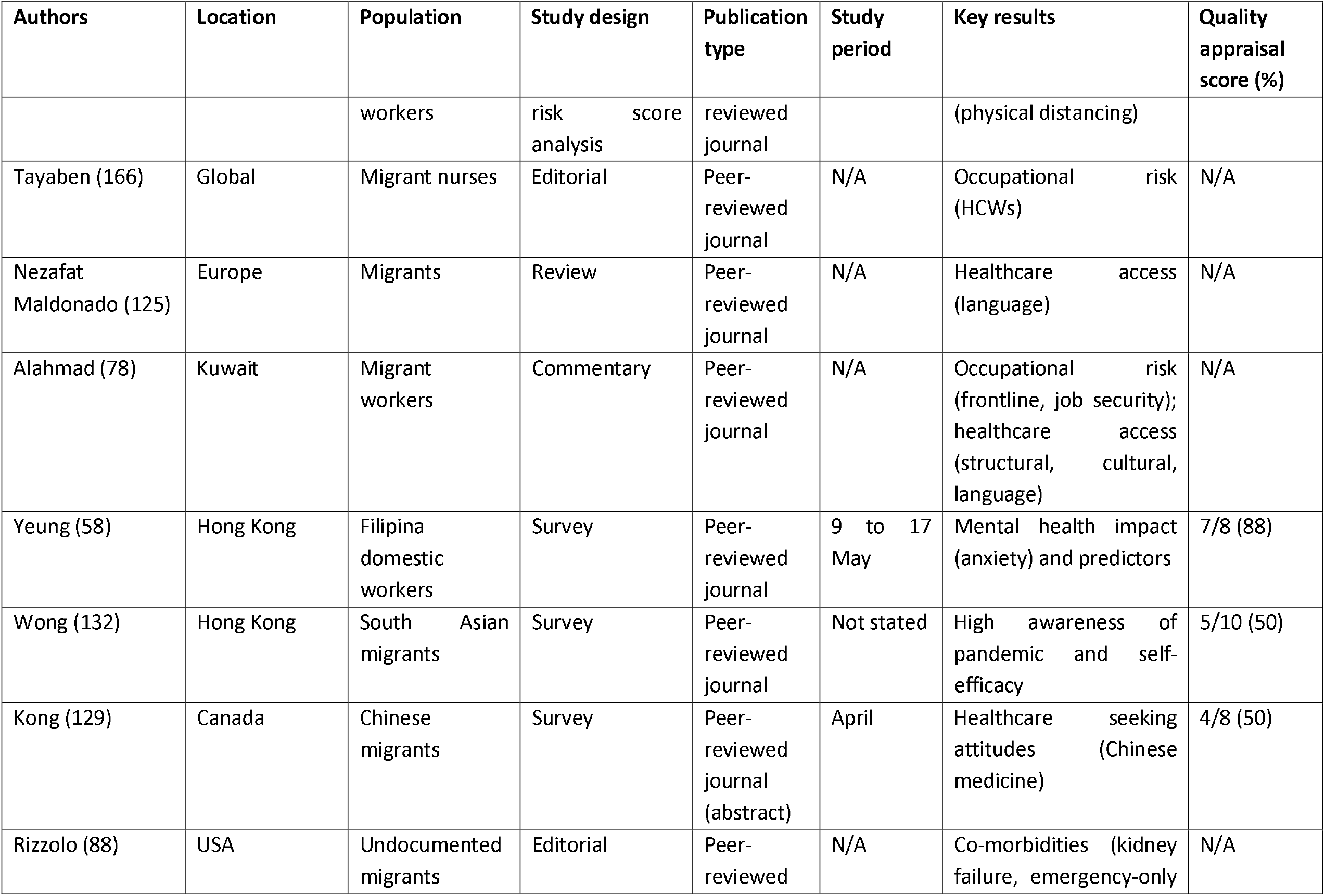
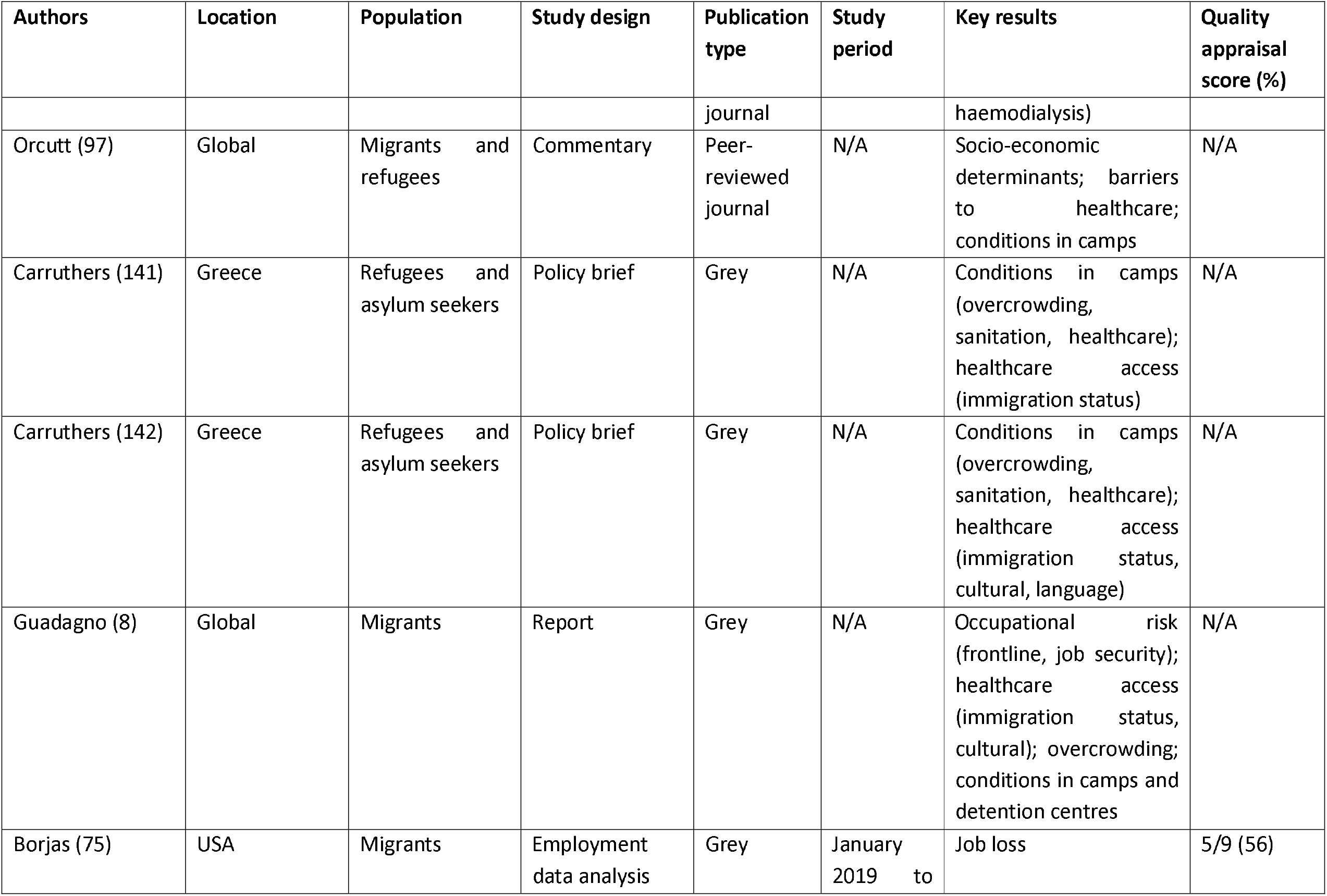
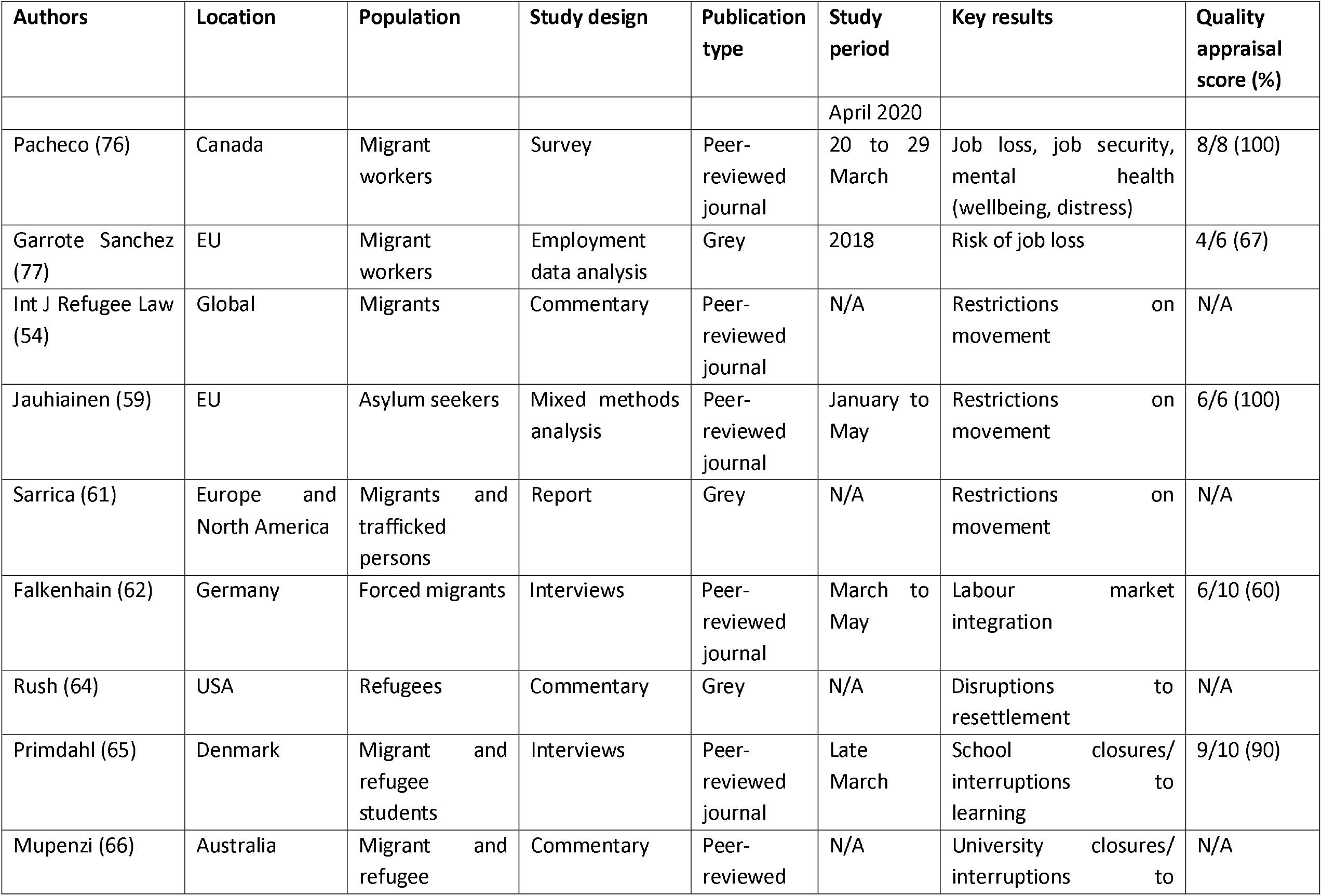
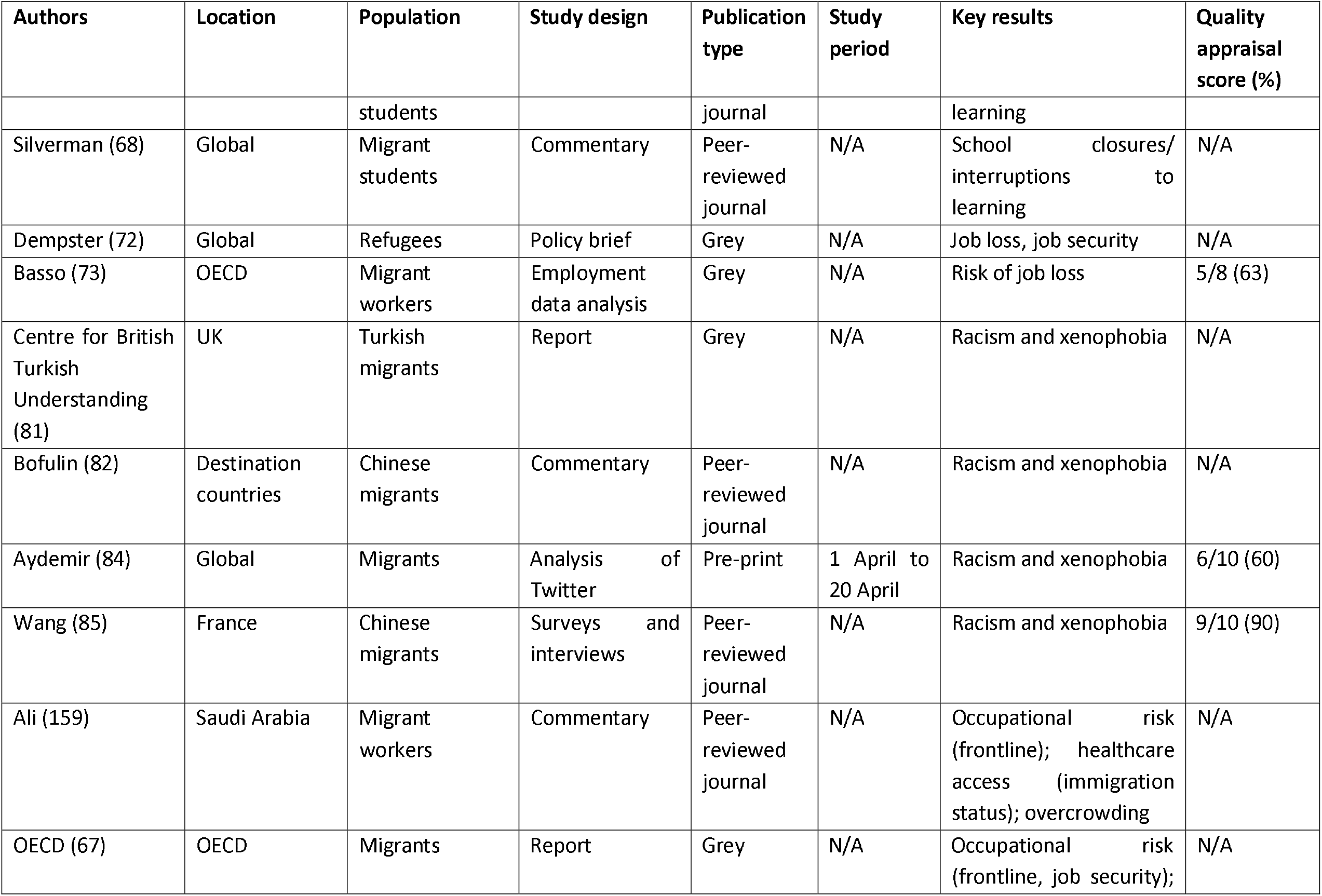
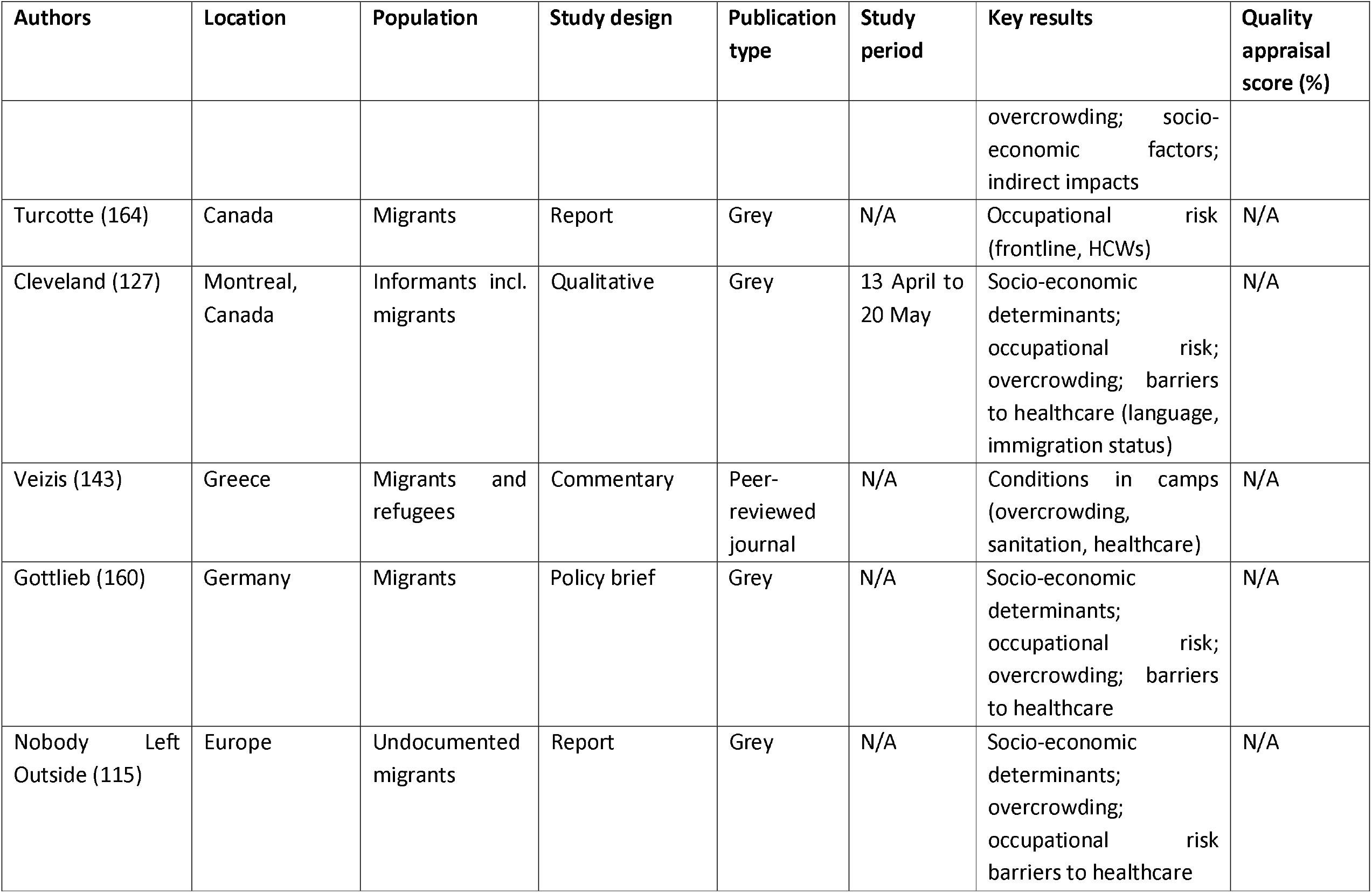
Characteristics of included data sources.

## Appendix 1: Search Strategy

TI:(Ancest* OR Diaspor* OR ethnic* OR Ethnoc* OR Ethnog* OR “Identity politics” OR Ingroups OR outgroups OR Intersectionality OR Kinship OR “Minority group*”∼3 OR “minority population*”∼2 OR minorities OR Multicultu* OR Polyethnic* OR “Population genetics” OR Race OR races OR racial OR Tribe* OR latino*) OR AB:(Ancest* OR Diaspor* OR ethnic* OR Ethnoc* OR Ethnog* OR “Identity politics” OR Ingroups OR Outgroups OR Intersectionality OR Kinship OR “Minority group*”∼3 OR “minority population*”∼2 OR minorities OR Multicultu* OR Polyethnic* OR “Population genetics” OR Race OR races OR racial OR Tribe* OR latino*) OR “afro american*”∼3 OR BAME OR latino* OR roma OR romani OR refugee* OR immigrant* OR “migrant” OR “displaced person” OR “displaced persons” OR “social determinant*”∼2 OR “latin population” OR “latin group*” OR “people of color” OR “people of colour”

## Appendix 2: World Bank High-Income Countries (2020)

1. Andorra
2. Antigua and Barbuda
3. Aruba
4. Australia
5. Austria
6. The Bahamas
7. Bahrain
8. Barbados
9. Belgium
10. Bermuda
11. British Virgin Islands
12. Brunei Darussalam
13. Canada
14. Cayman Islands
15. Channel Islands
16. Chile
17. Croatia
18. Curacao
19. Cyprus
20. Czech Republic
21. Denmark
22. Estonia
23. Faroe Islands
24. Finland
25. France
26. French Polynesia
27. Germany
28. Gibraltar
29. Greece
30. Greenland
31. Guam
32. Hong Kong SAR, China
33. Hungary
34. Iceland
35. Ireland
36. Isle of Man
37. Israel
38. Italy
39. Japan
40. Korea, Rep.
41. Kuwait
42. Latvia
43. Liechtenstein
44. Lithuania
45. Luxembourg
46. Macao SAR, China
47. Malta
48. Mauritius
49. Monaco
50. Nauru
51. Netherlands
52. New Caledonia
53. New Zealand
54. Northern Mariana Islands
55. Norway
56. Oman
57. Palau
58. Panama
59. Poland
60. Portugal
61. Puerto Rico
62. Qatar
63. Romania
64. San Marino
65. Saudi Arabia
66. Seychelles
67. Singapore
68. Sint Maarten (Dutch part)
69. Slovak Republic
70. Slovenia
71. Spain
72. St Kitts and Nevis
73. St Martin (French part)
74. Sweden
75. Switzerland
76. Trinidad and Tobago
77. Turks and Caicos Islands
78. United Arab Emirates
79. United Kingdom
80. United States
81. Uruguay
82. Virgin Islands

